# Dynamics of a dual SARS-CoV-2 strain co-infection on a prolonged viral shedding COVID-19 case: insights into clinical severity and disease duration

**DOI:** 10.1101/2020.12.22.20248392

**Authors:** Nicole Pedro, Cláudio N. Silva, Ana C. Magalhães, Bruno Cavadas, Ana M. Rocha, Ana C. Moreira, Maria S. Gomes, Diogo Silva, Joana Sobrinho-Simões, Angélica Ramos, Maria J. Cardoso, Rita Filipe, Pedro Palma, Filipa Ceia, Susana Silva, João T. Guimarães, António Sarmento, Verónica Fernandes, Luisa Pereira, Margarida Tavares

## Abstract

**Objectives:** A few molecularly proven SARS-CoV-2 cases of symptomatic reinfection are currently known worldwide, with a resolved first infection followed by a second infection after a 48 to 142-day intervening period. We report a multiple-component study of a clinically severe and prolonged viral shedding COVID-19 case in a teenager Portuguese female. She had two hospitalisations, a total of 19 RT-PCR tests, mostly positive, and criteria for releasing from home isolation at the end of 97 days.

**Methods:** The viral genome was sequenced in seven serial samples and in the diagnostic sample from an infected close relative. A human genome-wide array (>900K) was screened on the seven samples, and in vitro culture was conducted on isolates from three late samples.

**Results:** The patient had co-infection by two SARS-CoV-2 strains, affiliated in distinct clades and diverging by six variants. The 20A lineage was absolute at the diagnosis (shared with a cohabitating relative), but nine days later the 20B lineage had 3% frequency, and two months later the 20B lineage had 100% frequency. The 900K profiles confirmed the identity of the patient in the serial samples, and allowed us to infer that she had polygenic risk scores for hospitalization and severe respiratory disease within the normal distributions for a Portuguese population cohort.

**Conclusions:** The early-on dynamic co-infection was the probable cause for the severity of COVID-19 in this otherwise healthy young patient, and for her prolonged SARS-CoV-2 shedding profile.

## Introduction

The new Coronavirus disease 2019 (COVID-19), caused by the severe acute respiratory syndrome coronavirus 2 (SARS-CoV-2), was identified in Wuhan, China [1] and rapidly disseminated at a global scale. While the world is currently in the countdown for a vaccine, whether or not SARS-CoV-2 infection induces long-term protective immunity remains an open question. Closely related with the immunity issue is the puzzling observation of persistent viral shedding detectable in real time reverse transcriptase–polymerase chain reaction (RT-PCR) tests, even after symptom resolution [2]. This extended or recurrent positivity could be due to: (1) a reactivation of the virus after a period of clinical latency; (2) SARS-CoV-2 reinfection; (3) or simply RT-PCR tests detecting viral remains and not necessarily active viral particles. To untangle between the two first possibilities, it is necessary to perform viral whole-genome sequencing and detect genetically distinct strains of SARS-CoV-2 in each of the disease episodes; of course, reinfections by the same strain may remain unnoticed [3]. To check for the last possibility, the inclusion of infectivity studies might help understand if the virus retains both viability and integrity [4].

At the moment [5], a few molecularly proven SARS-CoV-2 symptomatic reinfections have been published worldwide, namely in Hong-Kong [6], Belgium [7], Ecuador [8] and USA [9]. After being considered recovered, these four patients presented a second infection by a genetically distinct SARS-CoV-2 strain after a 48 to 142-day intervening period. The latter two patients displayed a more severe disease in the second episode, in accordance to the hypothesis of an antibody-dependent enhancement (ADE) to SARS-CoV-2 [10]. The number of reinfection cases must be higher than currently reported as they are easily missed when asymptomatic. So far, two asymptomatic reinfections were detected in Indian healthcare workers, whom were routinely screened in their workplace [11]. Although these two cases were asymptomatic in the first and second infections, the viral load was higher in the second one, as inferred from the RT-PCR cycle threshold. Another theoretical possibility is the occurrence of reinfection while the first infection was not yet cured, a situation best described as co-infection. This hypothesis was advanced by the authors for the USA reinfection case [9], but not thoroughly studied.

In this work, we report the first molecularly proven dynamic early-on co-infection by two genetically distinct SARS-CoV-2 strains. This was a prolonged viral shedding case (97-day long), with a first severe disease manifestation, followed by a short-second hospitalisation episode, in an otherwise healthy young female.

## Methods

### RT-PCR and Antibody Testing

For detection of viral RNA by RT-PCR, each tube sample contained a nasopharyngeal and an oropharyngeal swab immersed in virus preservation solution. RNA was extracted with the Qiacube extractor by using the spin-column Qiamp virus minikit (Qiagen, Hilden, Germany). The reported RT-PCR results for the SARS-CoV-2 E gene were obtained with the LightCycler® Multiplex RNA Virus Master (Roche Life Science, Penzberg, Germany) at a LightCycler® 480 Instrument II (Roche Life Science Penzberg, Germany), and including a RNA extraction control (LightMix® Modular EAV RNA Extraction Control; Tib Molbiol, Berlin, Germany). Positive and negative controls were routinely included with each batch of tests. Relative quantification of the sample crossing points (cp) was automatically inferred with the LightCycler software.

The serological test used was a chemiluminescent microparticle immunoassay for qualitative detection of IgG against SARS-CoV-2 nucleoprotein (Abbott Diagnostics, Chicago, USA). Serum samples were run on the Abbott Architect instrument following the manufacturer’s instructions. The amount of IgG antibodies to SARS-CoV-2 in each sample was determined by comparing its chemiluminescent relative light unit (RLU) to the calibrator RLU (index S/C). A signal/cut-off (S/CO) ratio of ≥1.4 was interpreted as reactive and an S/CO ratio of <1.4 was interpreted as non-reactive.

The research protocol was approved by the Ethics Committee of the CHUSJ and University of Porto Medical School (process number: 102-20). The patient and her close relative provided written informed consent for publication.

### Viral Whole Genome-Sequencing and Phylogenetic Analysis

Seven serial extracted viral RNA samples from the patient and the diagnosis sample from her close relative were also used for the viral whole-genome sequencing. The protocol consisted in: reverse-transcription with the SuperScript™ VILO™ cDNA synthesis Kit (Thermo Fisher Scientific, Waltham, MA, USA); PCR enrichment of the SARS-CoV-2 genome and five human gene expression controls with the Ion AmpliSeq™ SARS-CoV-2 Research Panel; library construction with the Ion AmpliSeq™ Library Kit Plus; library quantification and size range verification at the 2200 TapeStation Automated Electrophoresis System, using the High Sensitivity DNA ScreenTape (Agilent Technologies, Santa Clara, CA, USA); next-generation sequencing (NGS) on the Ion S5XL system with the Ion 530™ chip; raw data extracted with the Ion Torrent pipeline. The bioinformatic pipeline consisted in: alignment of the raw data versus the reference genome (accession number NC_045512.2) with BWA tool; variant calling with three tools, FreeBayes, BCFtools and GATK, with editing of variants identified in at least two; variant annotation with SnpEff; consensus sequence was inferred with Bcftools [12]. Phylogenetic analysis and lineage/clade affiliation were done by merging the consensus sequences with publically available SARS-CoV-2 whole genomes from ViPR [13] with Mafft [14] (first 130bp and last 50bp were masked). The rooted phylogenetic tree was obtained with IQ-TREE 2 [15], and visualised using Interactive Tree Of Live version 4 [16]. The relative viral load in the samples (in molarity) was corrected from the TapeStation-based library quantification by removing the proportion of human reads (included in the panel and spurious ones).

### Genome-Wide Array Screening and Calculation of the Polygenic Risk Score

The genotyping of over 900,000 SNPs (900K) was attained with the Axiom™ Precision Medicine Diversity Array and the GeneTitan Multi-Channel (MC) Instrument (Thermo Fisher Scientific, Waltham, MA, USA). The laboratorial procedure was adapted to allow the use of samples extracted from the nasopharyngeal swabs, by increasing the time of the whole genome amplification step from 24h to 48h. Genotyping was inferred with Array Power Tool, and PADRE algorithm [17] was used for accurate estimate of shared ancestry (in this case, identity). These data were also used to calculate the polygenic risk score (PRS) in the Portuguese population cohort (n=198) to contextualise the patient score. The PMDA variants were uploaded into the Michigan Imputation Server (https://imputationserver.sph.umich.edu/index.html#!) and imputed based on the Haplotype Reference Consortium panel. The PRS values were calculated from the significant odd ratios (p-value<10^−5^) reported online by the COVID-19 host genetics initiative (https://www.covid19hg.org/) [18], for two phenotype cohorts from data released in 30^th^ September 2020 (COVID19-hg GWAS meta-analyses round 4 (alpha)): A2_ALL dataset of “very severe respiratory confirmed covid” (n=2,972) vs. population (n=284,472); and B2_ALL dataset of “hospitalised covid” (n=6,492) vs. population (n=1,012,809). Linked variants were removed in plink using the flags --clump-r2 0.4 --clump-kb 250; we ended up with 91 variants for the first dataset and 79 for the second. Plink was also used to estimate PRS values via an additive model, as the sum of the risk alleles, weighted by the effect size estimates from the genome-wide association study.

### In Vitro Culture of the Virus

To ascertain if the virus was still viable in the samples after the second hospitalization, in vitro culture in Vero cells (ATCC CCL-81; ATCC, Manassas, VA, USA) was performed, followed by SARS-CoV-2 spike antibody (GeneTex, Irvine, CA, USA) immunofluorescence detection. Images were acquired on the IN Cell Analyzer 2000 (Cytiva, Marlborough, MA, USA). Following a first inoculation of the samples for 96h, the resulting supernatant was transferred to 96 wells and inoculated for 24h, 48h and 72h in duplicates, to assess residual virus particles not detected in the first culture. We included a recently-diagnosed sample from another patient to guarantee the assay was able to detect the viral particles.

## Results

### Clinical features

A previously healthy young female presented to the local hospital emergency department in the beginning of March reporting a 9-day history of sustained fever, dry cough, pleuritic chest pain and vomiting. She was hemodynamically stable (105/63 mmHg blood pressure), but tachypnoeic (28 cpm respiratory rate), hypoxic to 88% on room air and febrile to 101.3°F. Her chest computed tomography (CT) scan revealed extensive bilateral subpleural ground-glass opacities (GGO) with areas of air-space consolidation, and she was admitted for aetiological investigation and treatment (Supplementary Figure S1). A nasopharyngeal swab performed during the initial workup detected SARS-CoV-2 RNA (Supplementary Table S1) and the patient was transferred to our referral centre for Emerging Infectious Diseases. At admission lymphopenia, a mild increased level of C-reactive protein and normal prothrombin and activated partial thromboplastin times were seen (Supplementary Table S2). Due to worsening respiratory status and increasing supplementary oxygen demands, she was placed on·High-Flow Nasal Oxygen (HFNO). After a 12-hour HFNO trial without improvement, she was admitted to our Infectious Diseases ICU on the 12^th^ day of symptoms, beginning an off-label 5-day hydroxychloroquine course.

After six days of inpatient care, she complained of left upper limb pain with signs consistent with deep vein thrombosis associated with indwelling peripheral venous catheter. She also complained of worsening bilateral pleuritic chest pain. Accordingly, D-dimer levels increased to 2.47 μg/mL and chest angio-CT scan showed lower left lung lobe peripheral infarction (Supplementary Figure S2). Anticoagulation with low-molecular-weight-heparin was started, the supplementary oxygen demands gradually decreased and she was discharged home 9 days after admission completely asymptomatic with an oxygen saturation of 99% on room air. RT-PCR tests of nasopharyngeal specimens remained positive for SARS-CoV-2 RNA at discharge. The patient was asked to continue isolating at home until attaining two consecutive negative tests.

Nearly two months after discharge, she was re-admitted with headaches, fever, myalgia and right pleuritic chest pain. Her vital signs were: 110/52 mmHg blood pressure, 112 bpm heart rate and 98.7°F body temperature. Her peripheral blood oxygen saturation was 97% on room air, and all of the blood work values were within the normal range, non-immunocompromised (Supplementary Tables S1 and S3). After an extensive microbiologic workup (Supplementary Table S2), other infectious aetiologies were ruled out. The chest CT scan (Supplementary Figure S3) revealed improved aeration of the lungs and resolving GGO features. Repeated SARS-CoV-2 RT-PCR was still positive. The symptoms resolved in 2 days. Anti-SARS-CoV-2 IgG antibodies were detected and reactive by immunoassay 77 days (relative light unit (RLU) index sample/calibrator (S/C) of 7.16), 91 days (index S/C of 6.89) and again 226 days after diagnosis, almost 8 months after first symptoms (index S/C of 2.26).

Only one household contact of the patient developed symptomatic COVID-19. In this case, the symptoms were mild, limited to a one-day low grade fever, and scarce dry cough starting 10 days after the patient’s symptom onset.

### Viral Load, Viability and Genome Analysis

The relative viral load on the nasopharyngeal samples along the disease course (Figure 1), inferred from the library quantification, showed a dramatic decrease (650 times lower) between the time of diagnosis and day nine when the patient was first discharged from the hospital. The viral load increased again by the 2^nd^ inpatient stay (inferred from the cp values) and was still relatively high two and three weeks after the second discharge (10-25 times higher than at first discharge). Nonetheless, it was impossible to retrieve integral and infectious SARS-CoV-2 viral particles from these samples through in vitro culture (Figure 2), despite some of these samples being still positive in the RT-PCR.

**Figure 1.**
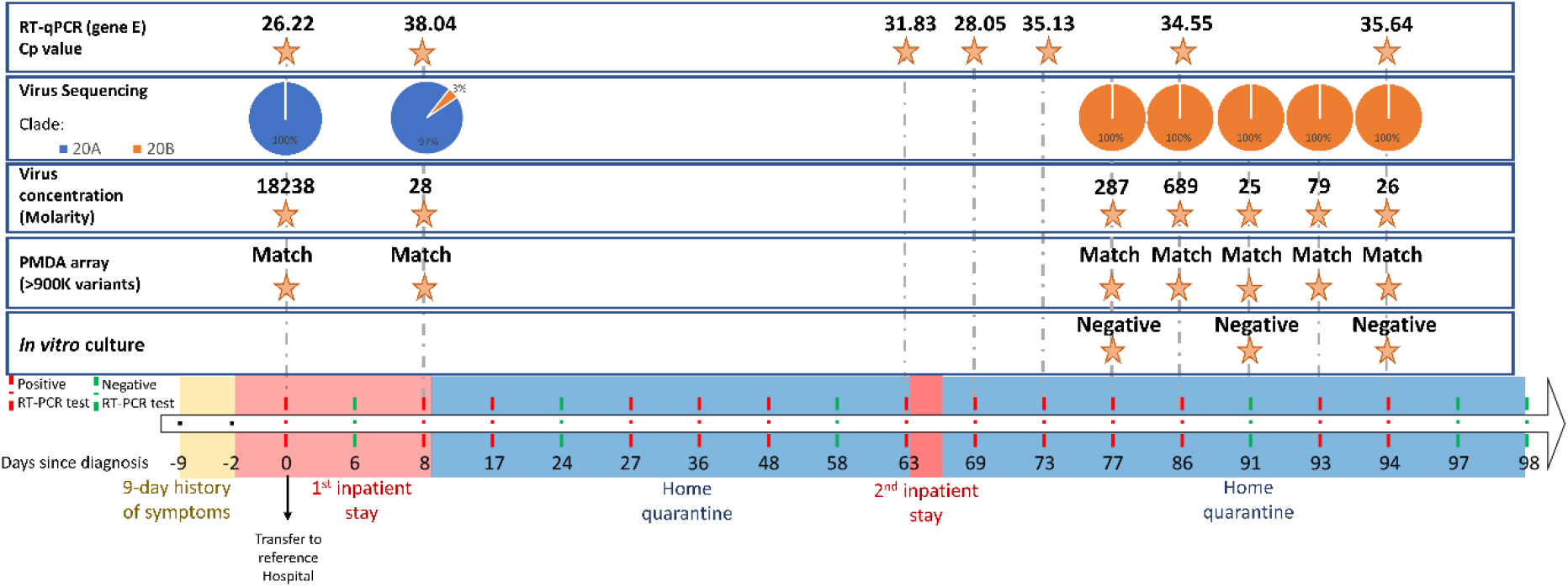
Timeline of the reported COVID-19 case and main results of the multi-component study conducted on the serial samples. The timeline bar indicates the periods of: symptom onset (yellow); inpatient stays (pink; the black arrow indicates the moment when the patient was transferred to the referral centre for Emerging Infectious Diseases); and, home quarantine (blue). It also includes the dates of all the 19 RT-PCR tests performed (green for negative result and red for positive result). The molecular tests were focused either on the virus (RT-PCR crossing point, proportion of clade/lineage affiliation inferred from viral whole-genome sequencing, virus concentration on the sequencing library, and in vitro culture) or the host genome-wide diversity (array containing >900K variants).

**Figure 2.**
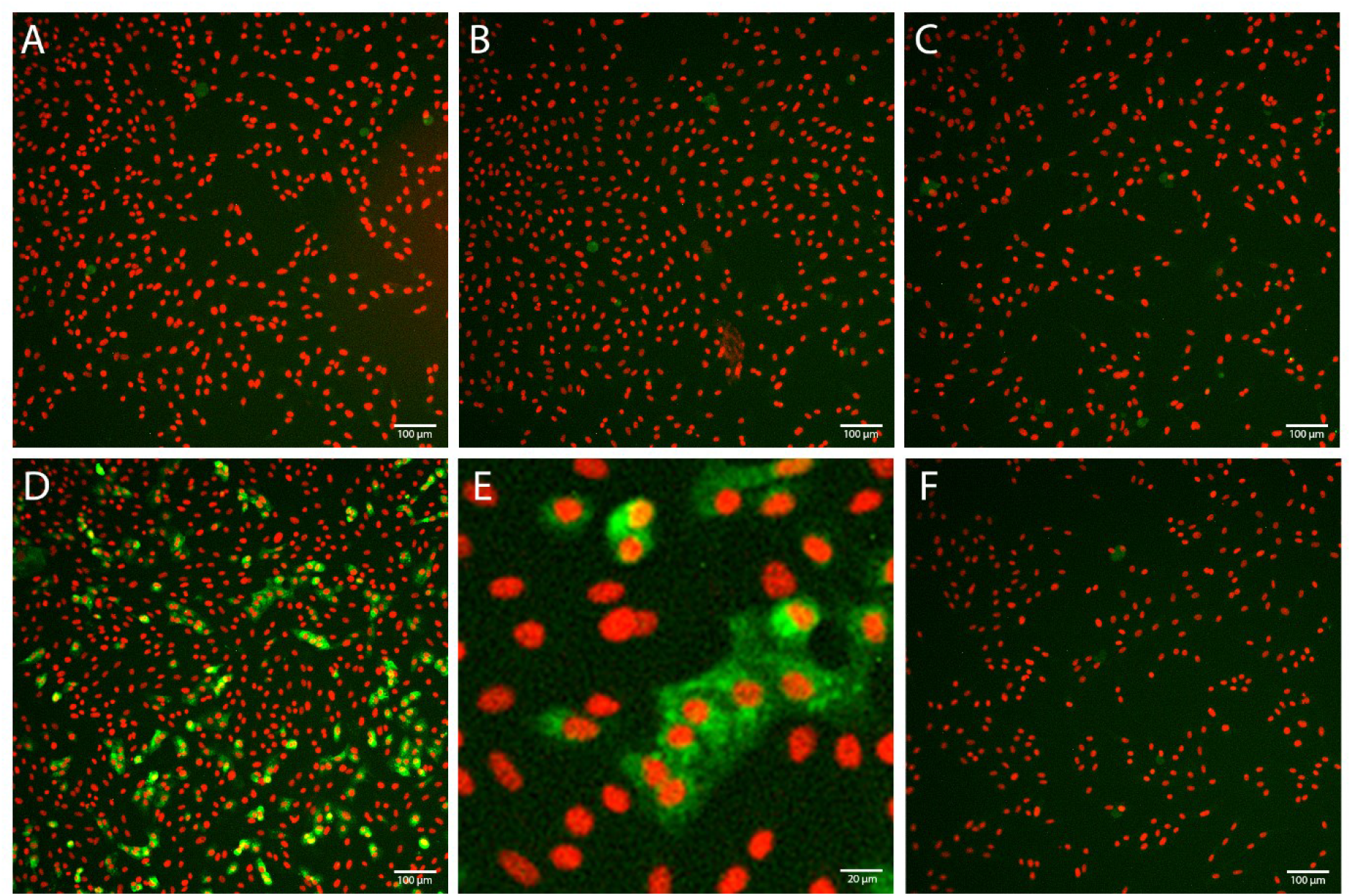
Results from the in vitro culture of SARS-CoV-2 from the nasopharyngeal samples. Images from immunofluorescence staining with the antibody against SARS-CoV-2 spike protein (green) and the marker for the nucleus (red), at 48h after the second inoculation in Vero cells. Images were acquired in an IN Cell Analyzer 2000, with 10x objective. Negative results in samples from the patient at 77 (A), 91 (B) and 94 (C) days after diagnosis. Positive result (D) and higher detail (E) for a recently-diagnosed sample from another patient. Negative result (F) for control of non-infected cells. Bars represent 100μm or 20μm.

The sequencing of the viral genome in the serial samples from the patient revealed very interesting results (Figure 1 and 3). At the diagnosis (sample P1.1), she displayed 100% of a 20A affiliated haplotype, bearing the basal C241T-C3037T-C14408T-A23403G variants (in relation to the reference sequence), and additional C3140T-G24077T-C295557 variants. The G24077T variant has been extensively observed in Lombardy, Italy (https://www.gisaid.org/) [19], while the C3140T-C295557 variants were highly observed in Portugal, by the time of this patient infections, especially so around her geographical region (reported online by the National Institute of Health Ricardo Jorge; https://insaflu.insa.pt/covid19/). This viral haplotype was also obtained from the RNA extracted from the diagnostic sample of the close relative. Transmission is supposed to have occurred from the patient to the close relative. Other cohabitants never presented symptoms, and three and half months later they had no serological evidence of past SARS-CoV-2 infection. However, her asymptomatic boyfriend showed reactivity to specific anti-SARS-CoV-2 IgG antibodies.

**Figure 3.**
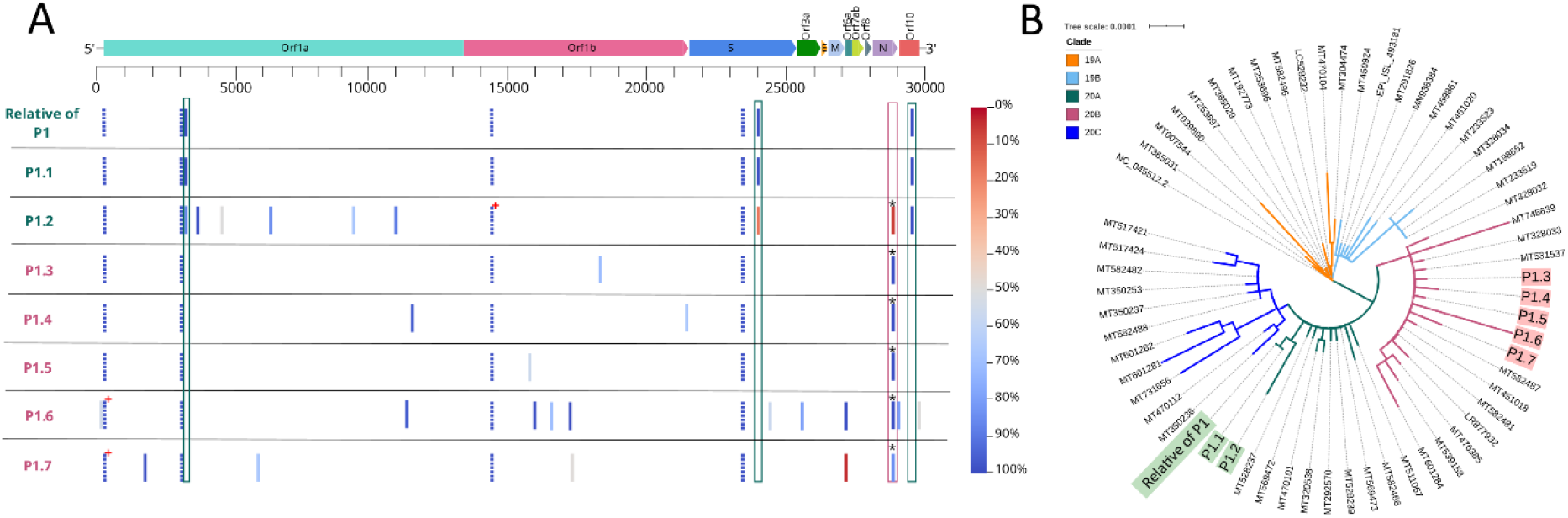
Detailed sequence diversity of the SARS-CoV-2 isolates from the various samples. A-Representation of the SARS-CoV-2 genome (genes are indicated) and the variants (only those with 50% frequency in at least one of the eight samples) sequenced in the isolates from the patient serial samples (P1.1 to P1.7) and her relative diagnostic sample. The colour code reflects the main 20A (in green) or 20B (in pink) SARS-CoV-2 strain. The gradient bar indicates the frequency of the variants. For easier visualization, the shared variants between 20A and 20B lineages are indicated in doted bars. The boxes highlight either the specific 20A variants (in green) or the 20B-defining variants (in pink; the asterisk calls the attention to the three sequential variants, G28881A-G28882A-G28883C). The red-cross indicates missing positions, matching known regions of SARS-CoV-2 genome that are difficult to sequence. B-Phylogenetic tree of the main SARS-CoV-2 clades known so far (19A, 19B, 20A 20B and 20C) and the sequences reported here (following the colour scheme of A).

**Figure 4.**
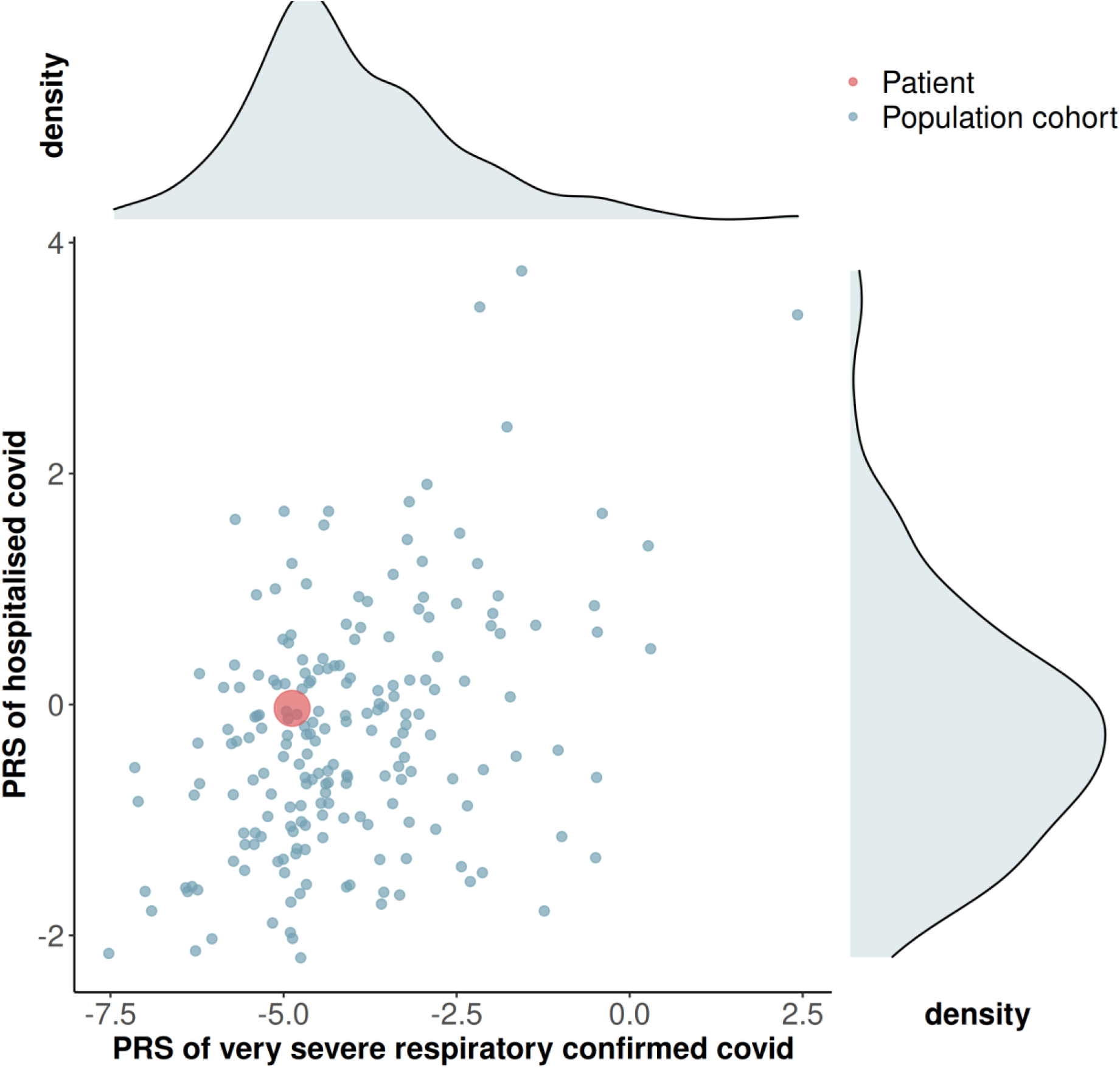
Scatter plot of the polygenic risk score (PRS) values and respective density plots for “very severe respiratory confirmed covid” and “hospitalised covid” phenotypes for the Portuguese population cohort (n=198), and the scatter point value for the patient (in red). The PRS values were calculated based on odd ratios reported online by the COVID-19 host genetics initiative (https://www.covid19hg.org/) [18].

The second serial sample (P1.2), at day nine after diagnosis and coinciding with the first hospital discharge, already revealed a mixed viral profile compatible with a 3% co-infection by a basal 20B affiliated haplotype. This 3%-20B haplotype is defined by G28881A-G28882A-G28883C variants, and shares C241T-C3037T-C14408T-A23403G variants with the 20A haplotype (hence, these shared variants had 100% frequency in P1.2). Thus, the two infecting haplotypes diverged by six SNPs. At least 12 days after the second hospital discharge (samples P1.3 to P1.7), the 20B lineage had totally replaced the initially dominant 20A lineage, including a sample with a negative RT-PCR result (P1.5). The samples with lower viral load (P1.2, P1.6 and P1.7) had other variants (Supplementary Table S4), at variable frequencies, in addition to the ones defining the lineages backbones. The variants that accumulate in the patient samples with lower viral load, or in other words, farther apart from the events that led to hospitalisations, can be due to two factors: (1) a lower resolution of our sequencing; (2) real representatives of the intra-host genomic diversity and plasticity of SARS-CoV-2 [20], a phenomenon known as quasispecies [21].

### Genome-Wide Array Screening and Polygenic Risk Score

We are certain that these RNA samples belong to the patient, and were not mismatched in the hospital or in the laboratories, through the characterization of an array containing 900K human variants. The 900K-profile was shared between the seven samples, and is unique of the patient, as she has no monozygotic twin. The 900K genotyping in the patient also allowed to estimate the PRS values for “very severe respiratory confirmed covid” and “hospitalised covid”, based on the incipient evidence that is being collected by the COVID-19 host genetics initiative [18]. The patient values were contextualised in a Portuguese population cohort (Figure 3; Supplementary Tables S5-S7). The PRS values for the patient were within the normal distributions observed in the population cohort, almost matching the mean values.

## Discussion

We illustrate a severe presentation of COVID-19 in a young healthy patient with prolonged viral shedding of SARS-CoV-2. This case was extremely challenging in terms of clinical diagnosis, and only the molecular study allowed us to shed light into its classification as the first proven SARS-CoV-2 co-infection.

The affiliation of the two haplotypes in distinct clades, which emerged at different time points along the pandemic (see https://nextstrain.org/sars-cov-2/) [22] renders it unlikely that they evolved intra-host from one-another. The co-infection with a virus belonging to a different clade had to occur in the 19-day period between beginning of symptoms and detection of the two strains. However, we hypothesize that co-infection was already present from disease onset, partially explaining the severe disease course of this healthy young female, and we simply failed to detect the second strain in the first diagnostic sample.

The episode that led to the second hospitalisation could be speculated as corresponding to the moment when the 20B strain became dominant. Unfortunately, we have no samples from this period allowing us to confirm this speculation. It is a fact that the viral load was higher (10-25 times) in the samples after this episode comparing with those from the date of the first discharge. But even so, we found no in vitro evidence that the patient was contagious at this period. These results agree with previous findings [23, 24] that a later-on positive RT-PCR does not imply the presence of active viral particles.

All this evidence seems to imply that the early-on co-infection by two SARS-CoV-2 strains was the cause of the severe disease displayed by this young and healthy female. Furthermore, the PRS evidence testified that she had no genetic predisposition for hospitalisation or severe COVID-19, when comparing with a Portuguese population cohort. The dynamics of exchange between dominant strains may have contributed to such prolonged viral shedding case. Most probably the close relative was only infected by one strain, hence displaying the milder disease, although we could not test it due to the unavailability of samples.

For sure, in the near future, several other cases of co-infection and reinfection will be identified as the pandemic continues. These cases will help to clarify if a worse disease course can be caused by the overlapping or sequential infection by different SARS-CoV-2 strains. The authors of the paper reporting the first reinfection in the USA patient [9], who presented with a second infection symptomatically more severe than the first, pointed to the possibility of co-infection instead of reinfection, implying that they simply did not detect overlapping “specimens A and B” in the April 2020 sample. In this case, as the patient was considered recovered after the first hospitalisation, he was not isolated, rendering the hypothesis of reinfection more probable. Reinfection would also agree with the more severe second COVID-19 episode presented by this patient. Both this case and the one we present here highlight the importance of performing more molecular studies on virus transmission dynamics. Either way, co-infection or reinfection, taking into account the possibility that the immunity driven by a specific SARS-CoV-2 strain does not protect against another strain, but can instead lead to a more severe disease pattern, is of extreme relevance in public health.

## Data Availability

Supplementary materials are available at Clinical Microbiology and Infection online. Extensive files listing the variant calls for the viral whole-genome sequences can be downloaded from https://portal.i3s.up.pt/docs/bcavadas/PEDROetal.zip. Given that the genome-wide profiles from the patient samples allow individual identification, researchers can request the data from the corresponding author, after justifying its need for the advancement of research.

## Acknowledgements

We wish to thank the patients who agreed to participate in this study. The authors acknowledge the support of the i3S Scientific Platforms BioSciences Screening and Genomics, members of the national infrastructure PPBI-Portuguese Platform of Bioimaging (PPBI-POCI-01-0145-FEDER-022122), PT-OPENSCREEN, GenomePT project (POCI-01-0145-FEDER-022184). And the authors also acknowledge the free information provided online by the COVID-19 host genetics initiative (https://www.covid19hg.org/).

## Contribution

NP performed the viral sequencing and array genotyping, with the technical supervision of AMR; BC established the bioinformatics pipelines, and together with NP and VF analysed the raw data; ACMagalhães, ACMoreira and MSG established the in vitro cultures and performed this assay with the patient samples; CNS and MT identified the patient case, contributed to clinical care and collected the clinical data; RF, PP, FC, SS and AS contributed to clinical care; DS, JSS and JTG were responsible for the RT-PCR diagnosis, while AR and MJC performed the serological tests, at the hospital environment; LP and MT conceptualised the study, and together with NP, VF and CNS wrote the manuscript that was later edited by all co-authors.

