## Supplemental table for "Dynamics of a dual SARS-CoV-2 strain co-infection on a prolonged viral shedding COVID-19 case: insights into clinical severity and disease duration"

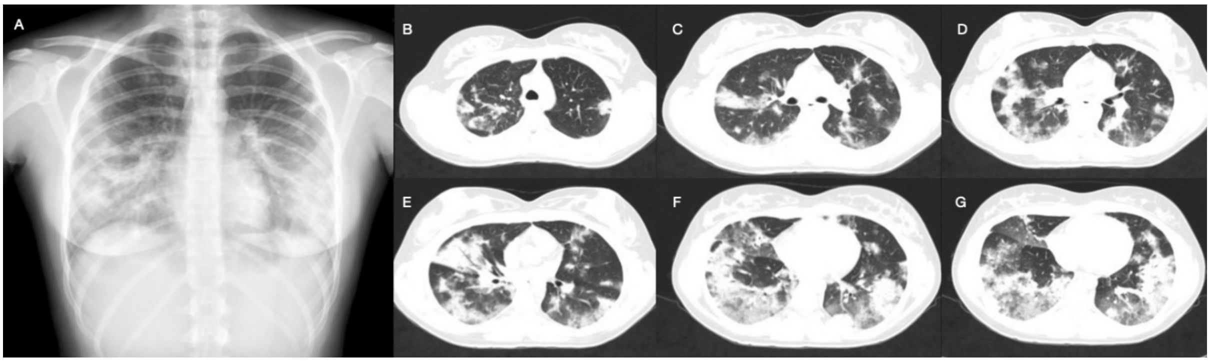


**Supplementary Figure S1.** X-ray **(A)** and chest CT-scan **(B-G)** pictures at the time of COVID-19 diagnosis revealing extensive bilateral subpleural ground-glass opacities (GGO) with areas of air-space consolidation concerning for COVID-19.

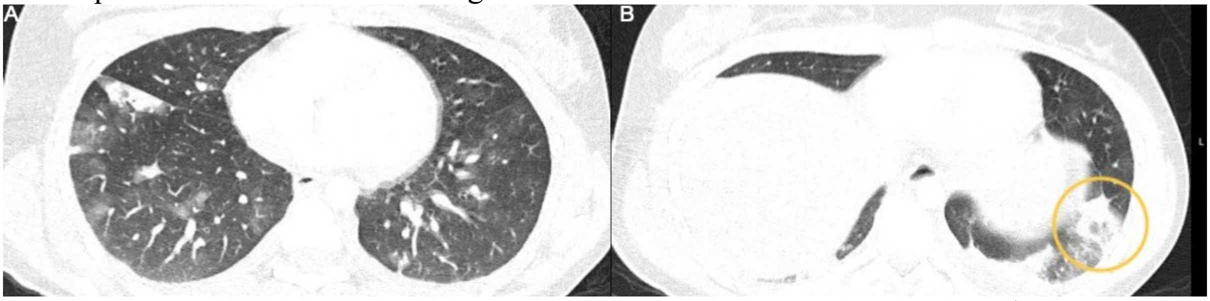


**Supplementary Figure S2.** Angio-chest CT scan performed after 6 days of 1st inpatient stay. Multiple foci of peripheral GGO and alveolar consolidation **(A)**. Heterogeneous ground-glass opacification with a peripheral halo of consolidation suggestive of pulmonary infarction of the left lung lobe (encircled in picture **B**).

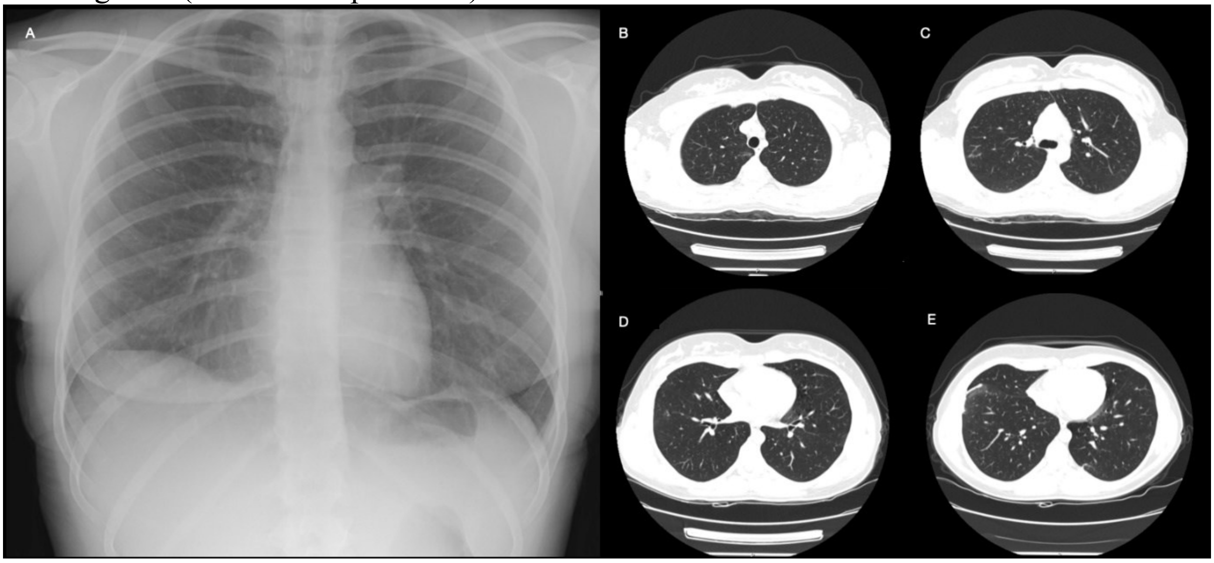


**Supplementary Figure S3.** X-ray **(A)** and chest CT-scan **(B-D)** pictures from re-admission, nearly 2 months after COVID-19 diagnosis, revealing improved aeration of the lung fields and resolving features of ground-glass-opacities.

**Supplementary Table S1. Results of the blood tests** collected at admission during the 1st inpatient stay and at the time of COVID-19 symptom recurrence that led to a 2nd inpatient stay. AST – Aspartate aminotransferase; ALT- Alanine Aminotransferase; GGT- Gamma-glutamyl-transferase; ALP– Alkaline Phosphatase; LDH – Lactate Dehydrogenase; CK – Creatinine Kinase; hs-cTNI – High-sensitivity Troponin I; CK-MB- Creatine Kinase MB; aPTT- Activated Partial Thromboplastin Time; PT- Prothrombin Time; CRP- C-reative-protein; NP- not performed.

| Blood tests | 1st inpatient stay results at admission | 2nd inpatient stay results at admission | Normal range values |
|---|---|---|---|
| **Haemoglobin (g/dL)** | 14.2 | 14.7 | 12.0-16.0 |
| **Leukocytes (x $10^9$/L)** | 5.7 | 6.7 | 4.0-11.0 |
| **Neutrophils (%)** | 82.6 | 58.2 | 53.8-69.8 |
| **Lymphocytes (%)** | 12.9 | 34.7 | 22.6-36.6 |
| **Platelets (x $10^9$/L)** | 172 | 290 | 150-400 |
| **AST (U/L)** | 34 | 19 | 10-31 |
| **ALT (U/L)** | 24 | 24 | 10-31 |
| **GGT (U/L)** | 41 | NP | 7-32 |
| **ALP (U/L)** | 50 | NP | 47-119 |
| **Total bilirubin (mg/dL)** | 0.60 | NP | < 1.20 |
| **Direct bilirubin (mg/dL)** | 0.16 | NP | < 0.40 |
| **LDH (U/L)** | 325 | NP | 135-225 |
| **CK (U/L)** | 74 | NP | 10-149 |
| **hs-cTnI (ng/L)** | < 1.9 | < 1.9 | < 16.0 |
| **CK-MB (ng/mL)** | 0.30 | 0.30 | 0.00-6.40 |
| **Mioglobin (ng/mL)** | 21.5 | 20.2 | < 146.9 |
| **Urea (mg/dL)** | 21 | 20 | 10-50 |
| **Plasma creatinine (mg/dL)** | 0.57 | 0.80 | 0.51-0.95 |
| **aPTT (sec)** | 27.2 | 30.7 | 24.2-36.4 |
| **PT (sec)** | 12.7 | 11.9 | 9.9-13.6 |
| **Fibrinogen (mg/dL)** | 382 | 248 | 200-400 |
| **CRP (mg/L)** | 37.6 | 0.40 | < 3.0 |
| **Ferritin (ng/mL)** | NP | 76.28 | 4.63-204 |

**Supplementary Table S2. Results of the microbiologic workup** collected at admission during the 1st inpatient stay and at the time of COVID-19 symptom recurrence that led to a 2nd inpatient stay. Ag – Antigen; NAAT – Nucleic Acid Amplification Test; NP- not performed.

| | Product | 1st inpatient stay results | 2nd inpatient stay results |
|---|---|---|---|
| Blood cultures (each pair) | Blood | negative | negative |
| Bacterial cultures | Sputum | negative | negative |
| Pneumococcal Ag | Urine | negative | negative |
| *Legionella* Ag | Urine | negative | negative |
| *Bordetella pertussis* NAAT | Naso/oropharyngeal swab | negative | negative |
| *Chlamydia pneumoniae* NAAT | Naso/oropharyngeal swab | negative | negative |
| *Mycoplasma pneumoniae* NAAT | Naso/oropharyngeal swab | negative | negative |
| Adenovirus NAAT | Naso/oropharyngeal swab | negative | negative |
| Coronavirus 229E, HKU1, NL63, OC43 NAAT | Naso/oropharyngeal swab | negative | negative |
| Metapneumovirus NAAT | Naso/oropharyngeal swab | negative | negative |
| Rhinovirus/Enterovirus NAAT | Naso/oropharyngeal swab | negative | negative |
| Influenza A & B NAAT | Naso/oropharyngeal swab | negative | NP |
| Parainfluenza 1,2,3 & 4 NAAT | Naso/oropharyngeal swab | negative | negative |
| Bocavirus NAAT | Naso/oropharyngeal swab | NP | negative |
| **SARS-CoV-2 NAAT** | **Naso/oropharyngeal swab** | **positive** | **positive** |

**Supplementary Table S3. Immunophenotyping** of peripheral blood lymphocytes and quantitative immunoglobulins' test performed during the 2nd inpatient stay.

| **Immunophenotyping of peripheral blood lymphocytes** | **Results** |
|---|---|
| **Lymphocytes (flow cytometry)** | 3059/mm$^3$ |
| **T populations:** | |
| CD3+ | 78.93% |
| CD3+CD4+ | 48.83% |
| (absolute value) | 1494/mm$^3$ |
| CD3+CD8+ | 27.19% |
| CD3+CD4+CD8+ | 0.33% |
| Ratio CD4+/CD8+ | 1.80 |
| **B populations:** | |
| CD19+ | 9.02% |
| **NK cell populations:** | |
| CD16&56+ | 11.62% |
| **Immunoglobulins** (normal range values) | **Results** |
| **Immunoglobulin G** (650-1500 mg/dL) | 955 |
| **IgG subclasses:** | |
| IgG1 (370.0-1280.0 mg/dL) | 705.0 |
| IgG2 (106.0-610.0 mg/dL) | 284.0 |
| IgG3 (18.0-163.0 mg/dL) | 42.6 |
| IgG4 (4.0-230.0 mg/dL) | 17.0 |
| **Immunoglobulin A** (78-312 mg/dL) | 137 |
| **Immunoglobulin M** (55-300 mg/dL) | 127 |

**Supplementary Table S4. List of variants** detected in all the analysed samples with a minimum of 5% frequency in at least one of the samples. The positions painted in blue are represented in Figure 3 and the grey coloured cells are the positions with variation.

| POS | REF | ALT | Relative of P1 | | P1.1 | | P1.2 | | P1.3 | | P1.4 | | P1.5 | | P1.6 | | P1.7 | |
|---|---|---|---|---|---|---|---|---|---|---|---|---|---|---|---|---|---|---|
| | | | N reads | Avg het | N reads | Avg het | N reads | Avg het | N reads | Avg het | N reads | Avg het | N reads | Avg het | N reads | Avg het | N reads | Avg het |
| **72** | G | A | 17979 | 0.0 | 33542 | 0.0 | 182 | 0.0 | 723 | 0.0 | 14180 | 0.0 | 74 | 0.0 | 942 | 54.6 | 20 | 0.0 |
| **128** | T | C | 13357 | 0.0 | 21817 | 0.0 | 7 | 0.0 | 20 | 0.0 | 2306 | 10.8 | 31 | 0.0 | 0 | 0.0 | 7 | 0.0 |
| **144** | T | C | 10366 | 0.0 | 20117 | 0.0 | 6 | 0.0 | 18 | 0.0 | 1999 | 11.8 | 24 | 0.0 | 0 | 0.0 | 6 | 0.0 |
| **186** | C | T | 11906 | 0.0 | 21162 | 0.0 | 8 | 0.0 | 18 | 0.0 | 2204 | 10.7 | 22 | 0.0 | 0 | 0.0 | 5 | 0.0 |
| **241** | C | T | 7627 | 100.0 | 16314 | 100.0 | 4 | 100.0 | 15 | 100.0 | 1422 | 100.0 | 4 | 100.0 | 0 | 0.0 | 0 | 0.0 |
| **262** | T | C | 11543 | 0.0 | 28796 | 0.0 | 13 | 0.0 | 29 | 0.0 | 3964 | 5.4 | 273 | 0.0 | 0 | 0.0 | 7 | 0.0 |
| **338** | G | A | 23813 | 0.0 | 43641 | 0.0 | 16 | 0.0 | 57 | 0.0 | 4718 | 6.0 | 13 | 0.0 | 1139 | 0.0 | 7 | 0.0 |
| **592** | T | C | 12027 | 0.0 | 38376 | 0.0 | 673 | 0.0 | 415 | 0.0 | 2560 | 9.0 | 408 | 0.0 | 0 | 0.0 | 22 | 0.0 |
| **621** | G | A | 12233 | 0.0 | 38804 | 0.0 | 679 | 0.0 | 414 | 0.0 | 2568 | 13.1 | 128 | 0.0 | 0 | 0.0 | 21 | 0.0 |
| **1204** | C | T | 8429 | 0.0 | 16516 | 0.0 | 6 | 0.0 | 43 | 0.0 | 3652 | 5.8 | 3 | 0.0 | 109 | 0.0 | 3 | 0.0 |
| **1367** | A | G | 5997 | 5.8 | 21891 | 0.0 | 9 | 0.0 | 39 | 0.0 | 4513 | 0.0 | 7 | 0.0 | 477 | 0.0 | 3 | 0.0 |
| **1459** | T | C | 10715 | 0.0 | 17352 | 0.0 | 7 | 42.9 | 32 | 0.0 | 3855 | 8.3 | 5 | 0.0 | 23 | 0.0 | 4 | 0.0 |
| **1745** | T | C | 5655 | 0.0 | 21029 | 0.0 | 4 | 0.0 | 18 | 0.0 | 563 | 0.0 | 1 | 0.0 | 2 | 0.0 | 2 | 100.0 |
| **1895** | G | A | 7627 | 0.0 | 26589 | 0.0 | 19 | 0.0 | 46 | 0.0 | 6302 | 0.0 | 6 | 0.0 | 0 | 0.0 | 9 | 33.3 |
| **2013** | C | T | 6990 | 0.0 | 30450 | 0.0 | 7 | 0.0 | 66 | 0.0 | 1349 | 11.8 | 11 | 0.0 | 295 | 0.0 | 66 | 0.0 |
| **2217** | T | C | 7873 | 0.0 | 29341 | 0.0 | 8 | 0.0 | 14 | 0.0 | 2895 | 6.4 | 15 | 0.0 | 18 | 0.0 | 11 | 0.0 |
| **2240** | A | G | 5786 | 0.0 | 27623 | 0.0 | 7 | 0.0 | 15 | 0.0 | 2489 | 7.3 | 13 | 0.0 | 13 | 0.0 | 10 | 0.0 |
| **2419** | A | G | 3638 | 0.0 | 15566 | 0.0 | 199 | 0.0 | 9 | 0.0 | 1410 | 8.5 | 10 | 0.0 | 12 | 0.0 | 6 | 0.0 |
| **2501** | A | G | 2556 | 0.0 | 13809 | 0.0 | 308 | 40.3 | 2 | 0.0 | 1413 | 0.0 | 4 | 0.0 | 75 | 0.0 | 9 | 0.0 |
| **2818** | A | G | 10039 | 0.0 | 36617 | 0.2 | 250 | 0.0 | 246 | 0.0 | 9009 | 0.0 | 9 | 0.0 | 1503 | 33.5 | 8 | 0.0 |
| **2901** | T | C | 16112 | 0.0 | 48138 | 0.3 | 134 | 5.2 | 1583 | 0.0 | 52328 | 2.0 | 26 | 0.0 | 65 | 0.0 | 25 | 0.0 |
| **2903** | A | G | 16016 | 0.0 | 47839 | 0.0 | 130 | 5.4 | 1595 | 0.0 | 52781 | 0.0 | 27 | 0.0 | 66 | 0.0 | 25 | 0.0 |
| **3037** | C | T | 10924 | 100.0 | 32832 | 100.0 | 201 | 100.0 | 1263 | 100.0 | 44978 | 100.0 | 5 | 100.0 | 3 | 100.0 | 18 | 100.0 |
| **3140** | C | T | 5925 | 99.9 | 19494 | 100.0 | 167 | 88.0 | 43 | 0.0 | 5805 | 0.0 | 2 | 0.0 | 1 | 0.0 | 26 | 0.0 |
| **3477** | T | C | 3917 | 0.0 | 8584 | 0.0 | 0 | 0.0 | 19 | 0.0 | 1365 | 5.4 | 1 | 0.0 | 1 | 0.0 | 0 | 0.0 |
| **3512** | A | G | 6594 | 0.0 | 17428 | 0.0 | 637 | 41.3 | 68 | 0.0 | 5492 | 0.0 | 6 | 0.0 | 1 | 0.0 | 8 | 0.0 |
| **3514** | G | A | 6561 | 0.0 | 17458 | 0.0 | 637 | 0.0 | 66 | 0.0 | 5525 | 7.3 | 5 | 0.0 | 1 | 0.0 | 8 | 0.0 |
| **3667** | T | C | 6776 | 0.0 | 20763 | 0.0 | 328 | 96.3 | 36 | 0.0 | 3371 | 0.0 | 5 | 0.0 | 83 | 0.0 | 6 | 0.0 |
| **4411** | A | G | 3685 | 0.0 | 18706 | 0.0 | 703 | 45.1 | 24 | 0.0 | 11 | 0.0 | 5 | 0.0 | 2 | 0.0 | 8 | 0.0 |
| **4535** | T | C | 7106 | 0.0 | 27151 | 0.3 | 181 | 0.0 | 34 | 0.0 | 6839 | 6.2 | 7 | 0.0 | 440 | 0.0 | 9 | 0.0 |
| **4543** | C | T | 9963 | 0.0 | 34131 | 0.0 | 230 | 54.8 | 54 | 0.0 | 8636 | 0.0 | 5 | 0.0 | 578 | 0.0 | 12 | 0.0 |
| **4672** | C | T | 1804 | 8.2 | 16600 | 0.0 | 5 | 0.0 | 12 | 0.0 | 7 | 0.0 | 3 | 0.0 | 1 | 0.0 | 5 | 0.0 |
| **4673** | A | G | 1320 | 6.1 | 15444 | 0.0 | 3 | 0.0 | 7 | 0.0 | 7 | 0.0 | 4 | 0.0 | 0 | 0.0 | 3 | 0.0 |
| **4759** | A | G | 6147 | 0.0 | 17337 | 0.0 | 70 | 0.0 | 148 | 0.0 | 7806 | 6.9 | 6 | 0.0 | 0 | 0.0 | 121 | 0.0 |
| **4956** | A | G | 6053 | 0.0 | 25312 | 0.0 | 307 | 0.0 | 66 | 0.0 | 2935 | 8.0 | 17 | 0.0 | 195 | 0.0 | 54 | 0.0 |
| **5030** | A | G | 5493 | 0.0 | 14940 | 0.0 | 341 | 0.0 | 37 | 0.0 | 3150 | 0.0 | 14 | 0.0 | 213 | 7.0 | 10 | 0.0 |
| **5055** | C | T | 5586 | 0.0 | 14608 | 0.0 | 339 | 0.0 | 35 | 0.0 | 2471 | 17.6 | 2 | 0.0 | 221 | 0.0 | 5 | 0.0 |

| POS | REF | ALT | Relative of P1 | | P1.1 | | P1.2 | | P1.3 | | P1.4 | | P1.5 | | P1.6 | | P1.7 | |
|---|---|---|---|---|---|---|---|---|---|---|---|---|---|---|---|---|---|---|
| | | | N reads | Avg het | N reads | Avg het | N reads | Avg het | N reads | Avg het | N reads | Avg het | N reads | Avg het | N reads | Avg het | N reads | Avg het |
| **5199** | G | A | 3703 | 0.0 | 12476 | 0.0 | 325 | 0.0 | 21 | 0.0 | 1895 | 11.4 | 1 | 0.0 | 2 | 0.0 | 2 | 0.0 |
| **5325** | A | G | 3861 | 0.0 | 9117 | 0.0 | 319 | 0.0 | 16 | 0.0 | 2044 | 7.5 | 0 | 0.0 | 25 | 0.0 | 1 | 0.0 |
| **5896** | A | G | 4374 | 0.0 | 16370 | 0.0 | 6 | 0.0 | 12 | 0.0 | 3013 | 0.0 | 2 | 0.0 | 29 | 0.0 | 3 | 66.7 |
| **5921** | G | A | 6482 | 0.0 | 18894 | 0.0 | 10 | 0.0 | 28 | 0.0 | 3745 | 0.0 | 0 | 0.0 | 35 | 28.6 | 4 | 0.0 |
| **5984** | T | A | 421 | 6.4 | 8061 | 0.0 | 0 | 0.0 | 4 | 0.0 | 233 | 0.0 | 1 | 0.0 | 0 | 0.0 | 4 | 0.0 |
| **6170** | G | A | 8993 | 0.0 | 30990 | 0.0 | 54 | 0.0 | 27 | 0.0 | 1652 | 11.1 | 6 | 0.0 | 258 | 0.0 | 10 | 0.0 |
| **6198** | C | T | 9860 | 0.0 | 32941 | 0.0 | 56 | 0.0 | 27 | 0.0 | 1821 | 0.0 | 6 | 0.0 | 262 | 5.7 | 11 | 0.0 |
| **6200** | T | C | 10871 | 0.0 | 34092 | 0.0 | 59 | 0.0 | 29 | 0.0 | 1950 | 9.4 | 7 | 0.0 | 273 | 0.0 | 11 | 0.0 |
| **6238** | T | C | 11250 | 0.0 | 34647 | 0.0 | 56 | 85.7 | 31 | 0.0 | 2016 | 0.0 | 9 | 0.0 | 284 | 0.0 | 12 | 0.0 |
| **6254** | G | A | 773 | 5.4 | 13265 | 0.0 | 55 | 0.0 | 12 | 0.0 | 164 | 0.0 | 6 | 0.0 | 292 | 0.0 | 8 | 0.0 |
| **6475** | A | G | 8374 | 0.0 | 23484 | 0.0 | 381 | 0.0 | 46 | 0.0 | 1536 | 12.4 | 17 | 0.0 | 0 | 0.0 | 9 | 0.0 |
| **6675** | C | T | 6242 | 0.0 | 31757 | 0.0 | 414 | 0.0 | 472 | 0.0 | 14436 | 12.4 | 12 | 0.0 | 1156 | 0.0 | 304 | 0.0 |
| **6701** | C | T | 6032 | 0.0 | 31314 | 0.0 | 407 | 0.0 | 463 | 0.0 | 14034 | 2.6 | 12 | 0.0 | 1113 | 11.8 | 313 | 0.0 |
| **6755** | G | A | 2079 | 0.0 | 18680 | 11.5 | 273 | 0.0 | 92 | 0.0 | 1742 | 0.0 | 7 | 0.0 | 566 | 0.0 | 279 | 0.0 |
| **6882** | T | C | 3205 | 0.0 | 8204 | 0.0 | 91 | 0.0 | 3 | 0.0 | 385 | 5.5 | 0 | 0.0 | 0 | 0.0 | 0 | 0.0 |
| **6920** | A | G | 3079 | 0.0 | 14260 | 0.0 | 166 | 0.0 | 10 | 0.0 | 370 | 9.5 | 4 | 0.0 | 0 | 0.0 | 3 | 0.0 |
| **6958** | T | C | 2343 | 0.0 | 14369 | 0.0 | 159 | 0.0 | 10 | 0.0 | 337 | 11.9 | 5 | 0.0 | 0 | 0.0 | 5 | 0.0 |
| **6982** | C | T | 2374 | 0.0 | 14384 | 0.3 | 153 | 0.0 | 8 | 0.0 | 327 | 8.0 | 5 | 0.0 | 0 | 0.0 | 5 | 0.0 |
| **6991** | T | C | 1657 | 0.0 | 12657 | 0.0 | 122 | 0.0 | 8 | 0.0 | 265 | 6.4 | 5 | 0.0 | 0 | 0.0 | 3 | 0.0 |
| **7225** | T | C | 1586 | 10.6 | 10328 | 0.6 | 21 | 0.0 | 4 | 0.0 | 737 | 0.0 | 1 | 0.0 | 0 | 0.0 | 1 | 0.0 |
| **7303** | C | T | 311 | 0.0 | 7085 | 12.1 | 43 | 0.0 | 1 | 0.0 | 0 | 0.0 | 4 | 0.0 | 0 | 0.0 | 4 | 0.0 |
| **7359** | T | C | 4327 | 0.0 | 17263 | 0.0 | 50 | 0.0 | 15 | 0.0 | 2459 | 0.0 | 12 | 33.3 | 0 | 0.0 | 26 | 19.2 |
| **7513** | T | C | 4487 | 0.0 | 19847 | 0.0 | 12 | 0.0 | 26 | 0.0 | 3676 | 5.1 | 5 | 0.0 | 1 | 0.0 | 18 | 0.0 |
| **7537** | A | G | 3878 | 0.0 | 19631 | 0.0 | 11 | 0.0 | 20 | 0.0 | 3474 | 5.0 | 3 | 0.0 | 1 | 0.0 | 13 | 0.0 |
| **8193** | A | G | 3485 | 7.3 | 15758 | 0.0 | 8 | 0.0 | 14 | 0.0 | 3050 | 0.0 | 5 | 0.0 | 246 | 0.0 | 9 | 0.0 |
| **8388** | A | G | 4947 | 0.0 | 18886 | 0.0 | 525 | 41.7 | 522 | 0.0 | 24309 | 0.0 | 9 | 0.0 | 1050 | 0.0 | 9 | 0.0 |
| **8389** | C | T | 4877 | 5.7 | 18899 | 0.0 | 511 | 0.0 | 529 | 0.0 | 23956 | 0.0 | 8 | 0.0 | 1008 | 0.0 | 9 | 0.0 |
| **8750** | A | G | 4464 | 3.9 | 19109 | 0.0 | 371 | 0.0 | 92 | 0.0 | 9222 | 0.0 | 8 | 0.0 | 388 | 5.7 | 6 | 0.0 |
| **8928** | T | G | 2626 | 4.7 | 12005 | 0.0 | 153 | 5.9 | 20 | 0.0 | 3965 | 2.3 | 3 | 0.0 | 259 | 0.0 | 3 | 0.0 |
| **9071** | A | G | 8749 | 0.0 | 25287 | 0.0 | 37 | 0.0 | 897 | 0.0 | 21944 | 0.4 | 5 | 0.0 | 831 | 34.1 | 21 | 0.0 |
| **9481** | T | C | 2743 | 0.0 | 13519 | 0.0 | 3 | 66.7 | 28 | 0.0 | 191 | 0.0 | 11 | 0.0 | 718 | 0.0 | 12 | 0.0 |
| **9704** | T | C | 2870 | 0.0 | 14830 | 0.0 | 2 | 0.0 | 8 | 0.0 | 473 | 5.9 | 5 | 0.0 | 0 | 0.0 | 2 | 0.0 |
| **10024** | A | G | 9789 | 0.0 | 27393 | 0.0 | 215 | 0.0 | 88 | 0.0 | 8892 | 0.0 | 2 | 0.0 | 1377 | 24.6 | 24 | 0.0 |
| **10201** | G | A | 7377 | 0.0 | 30149 | 0.0 | 169 | 0.0 | 78 | 0.0 | 7613 | 5.2 | 16 | 0.0 | 2 | 0.0 | 8 | 0.0 |
| **10389** | T | C | 4577 | 0.0 | 26143 | 0.0 | 616 | 0.0 | 84 | 0.0 | 11059 | 0.0 | 11 | 0.0 | 462 | 47.8 | 32 | 0.0 |
| **10409** | A | G | 4214 | 0.0 | 27736 | 0.0 | 589 | 0.0 | 70 | 0.0 | 10855 | 11.2 | 7 | 0.0 | 426 | 0.0 | 32 | 0.0 |
| **10411** | T | C | 5048 | 0.0 | 28279 | 0.0 | 625 | 0.0 | 88 | 0.0 | 11313 | 16.0 | 10 | 0.0 | 487 | 0.0 | 32 | 0.0 |
| **10549** | G | A | 4954 | 0.0 | 24894 | 0.0 | 284 | 0.0 | 29 | 0.0 | 3900 | 6.7 | 3 | 0.0 | 637 | 0.0 | 5 | 0.0 |
| **10622** | A | C | 1942 | 0.0 | 9178 | 0.0 | 81 | 9.9 | 20 | 0.0 | 50 | 0.0 | 2 | 0.0 | 277 | 0.0 | 7 | 0.0 |
| **10729** | A | G | 3480 | 0.0 | 25543 | 0.0 | 156 | 0.0 | 54 | 0.0 | 3701 | 6.3 | 6 | 0.0 | 1 | 0.0 | 13 | 0.0 |
| **10763** | T | C | 4324 | 0.0 | 25689 | 0.0 | 168 | 0.0 | 81 | 0.0 | 4000 | 8.4 | 8 | 0.0 | 1 | 0.0 | 19 | 0.0 |

| POS | REF | ALT | Relative of P1 | | P1.1 | | P1.2 | | P1.3 | | P1.4 | | P1.5 | | P1.6 | | P1.7 | |
|---|---|---|---|---|---|---|---|---|---|---|---|---|---|---|---|---|---|---|
| | | | N reads | Avg het | N reads | Avg het | N reads | Avg het | N reads | Avg het | N reads | Avg het | N reads | Avg het | N reads | Avg het | N reads | Avg het |
| **10973** | A | G | 3434 | 0.0 | 11312 | 0.0 | 270 | 9.6 | 15 | 0.0 | 5189 | 0.0 | 1 | 0.0 | 1 | 0.0 | 2 | 0.0 |
| **11076** | T | C | 4112 | 6.1 | 17403 | 0.0 | 273 | 0.0 | 15 | 0.0 | 4121 | 0.0 | 1 | 0.0 | 1 | 0.0 | 3 | 0.0 |
| **11083** | G | T | 4935 | 2.6 | 24865 | 0.9 | 119 | 89.9 | 18 | 0.0 | 5368 | 0.0 | 1 | 0.0 | 2 | 0.0 | 3 | 0.0 |
| **11180** | T | C | 3694 | 0.0 | 14495 | 0.0 | 143 | 0.0 | 28 | 0.0 | 1430 | 15.1 | 1 | 0.0 | 0 | 0.0 | 2 | 0.0 |
| **11185** | G | A | 6954 | 0.0 | 26870 | 0.0 | 259 | 0.0 | 45 | 0.0 | 2553 | 0.0 | 5 | 0.0 | 3 | 100.0 | 6 | 0.0 |
| **11243** | G | A | 4602 | 0.0 | 12203 | 0.0 | 196 | 0.0 | 29 | 0.0 | 2019 | 12.5 | 0 | 0.0 | 0 | 0.0 | 3 | 0.0 |
| **11454** | C | T | 5221 | 0.0 | 20942 | 0.0 | 48 | 0.0 | 36 | 0.0 | 3112 | 0.0 | 9 | 0.0 | 712 | 15.3 | 9 | 0.0 |
| **11638** | T | C | 2138 | 0.0 | 15689 | 0.0 | 164 | 0.0 | 22 | 0.0 | 316 | 100.0 | 4 | 0.0 | 0 | 0.0 | 16 | 0.0 |
| **11827** | A | G | 13786 | 0.0 | 56716 | 0.0 | 20 | 15.0 | 144 | 0.0 | 10927 | 5.7 | 8 | 0.0 | 2799 | 0.0 | 14 | 0.0 |
| **11851** | G | A | 17115 | 0.0 | 58806 | 0.0 | 26 | 19.2 | 182 | 0.0 | 12998 | 19.1 | 6 | 0.0 | 3057 | 0.0 | 17 | 0.0 |
| **11968** | T | C | 10865 | 0.0 | 42559 | 0.0 | 17 | 0.0 | 109 | 0.0 | 5742 | 0.0 | 8 | 0.0 | 2463 | 15.7 | 18 | 0.0 |
| **12519** | A | G | 7656 | 0.0 | 27051 | 0.0 | 371 | 0.0 | 100 | 0.0 | 3789 | 0.0 | 12 | 33.3 | 0 | 0.0 | 8 | 0.0 |
| **12613** | G | A | 9133 | 0.0 | 41789 | 0.0 | 86 | 0.0 | 142 | 0.0 | 3083 | 6.5 | 21 | 0.0 | 1 | 0.0 | 23 | 0.0 |
| **12652** | A | G | 9938 | 0.0 | 42570 | 0.0 | 90 | 0.0 | 158 | 0.0 | 3157 | 5.9 | 25 | 0.0 | 0 | 0.0 | 24 | 0.0 |
| **12670** | T | C | 10092 | 0.0 | 42207 | 0.0 | 92 | 0.0 | 155 | 0.0 | 3168 | 0.0 | 23 | 17.4 | 1 | 0.0 | 23 | 0.0 |
| **13401** | A | G | 9064 | 0.0 | 20811 | 0.0 | 416 | 0.0 | 102 | 0.0 | 4535 | 5.1 | 46 | 0.0 | 283 | 0.0 | 278 | 0.0 |
| **13415** | G | A | 9137 | 0.0 | 21132 | 0.0 | 413 | 0.0 | 105 | 0.0 | 4597 | 0.0 | 49 | 0.0 | 291 | 0.0 | 282 | 9.9 |
| **13427** | G | A | 9811 | 0.0 | 21147 | 0.0 | 420 | 0.0 | 105 | 0.0 | 4712 | 0.0 | 48 | 0.0 | 284 | 6.7 | 280 | 0.0 |
| **13470** | G | A | 8206 | 0.0 | 17507 | 0.0 | 309 | 0.0 | 11 | 0.0 | 3583 | 6.3 | 5 | 0.0 | 0 | 0.0 | 0 | 0.0 |
| **13616** | A | G | 3452 | 0.0 | 12825 | 0.0 | 0 | 0.0 | 7 | 0.0 | 779 | 12.7 | 0 | 0.0 | 0 | 0.0 | 2 | 0.0 |
| **13875** | A | G | 8103 | 0.0 | 24955 | 0.0 | 10 | 0.0 | 27 | 0.0 | 1192 | 16.4 | 5 | 0.0 | 228 | 0.0 | 3 | 0.0 |
| **14006** | C | A | 8841 | 0.0 | 33679 | 0.0 | 12 | 0.0 | 77 | 0.0 | 3682 | 5.0 | 11 | 0.0 | 703 | 0.0 | 7 | 0.0 |
| **14188** | G | A | 6957 | 6.2 | 27983 | 0.0 | 6 | 0.0 | 45 | 0.0 | 3369 | 0.0 | 12 | 0.0 | 254 | 0.0 | 13 | 0.0 |
| **14408** | C | T | 445 | 100.0 | 9840 | 100.0 | 0 | 0.0 | 3 | 66.7 | 86 | 0.0 | 187 | 100.0 | 921 | 100.0 | 3 | 100.0 |
| **14424** | A | G | 984 | 0.0 | 10483 | 0.0 | 26 | 0.0 | 186 | 0.0 | 10411 | 7.3 | 191 | 0.0 | 942 | 0.0 | 4 | 0.0 |
| **14530** | T | C | 2463 | 5.3 | 14456 | 0.0 | 45 | 0.0 | 259 | 0.0 | 17223 | 0.0 | 5 | 0.0 | 1033 | 0.0 | 20 | 0.0 |
| **14586** | T | C | 2009 | 0.0 | 14203 | 0.0 | 1 | 0.0 | 41 | 0.0 | 1 | 0.0 | 6 | 0.0 | 746 | 0.0 | 20 | 40.0 |
| **14846** | T | C | 5465 | 0.0 | 18377 | 0.0 | 135 | 14.1 | 18 | 0.0 | 2702 | 0.0 | 5 | 0.0 | 0 | 0.0 | 8 | 0.0 |
| **14857** | G | A | 4915 | 0.0 | 18510 | 0.0 | 136 | 5.2 | 15 | 0.0 | 2587 | 0.0 | 6 | 0.0 | 0 | 0.0 | 8 | 0.0 |
| **14899** | T | C | 1033 | 0.0 | 7699 | 0.0 | 140 | 0.0 | 14 | 0.0 | 43 | 39.5 | 2 | 0.0 | 0 | 0.0 | 4 | 0.0 |
| **14913** | C | T | 994 | 0.0 | 7670 | 0.0 | 134 | 0.0 | 24 | 33.3 | 336 | 0.0 | 1 | 0.0 | 0 | 0.0 | 3 | 0.0 |
| **15189** | A | G | 13945 | 0.0 | 36581 | 0.2 | 299 | 0.0 | 63 | 0.0 | 5393 | 0.0 | 19 | 0.0 | 312 | 0.0 | 15 | 40.0 |
| **15201** | A | G | 14150 | 0.0 | 36618 | 0.0 | 300 | 0.0 | 61 | 0.0 | 5381 | 0.0 | 17 | 0.0 | 322 | 0.0 | 15 | 33.3 |
| **15444** | G | A | 24934 | 0.0 | 32766 | 0.0 | 486 | 11.1 | 390 | 0.0 | 15905 | 0.0 | 16 | 0.0 | 0 | 0.0 | 10 | 0.0 |
| **15570** | T | C | 30872 | 0.0 | 62167 | 0.0 | 484 | 10.3 | 694 | 0.0 | 16112 | 0.0 | 237 | 0.0 | 1389 | 0.0 | 24 | 0.0 |
| **15725** | C | T | 18173 | 0.0 | 40821 | 0.0 | 10 | 0.0 | 364 | 0.0 | 3109 | 0.0 | 264 | 0.0 | 1482 | 0.0 | 22 | 13.6 |
| **15758** | A | G | 9238 | 0.0 | 10576 | 0.0 | 8 | 0.0 | 9 | 0.0 | 1871 | 7.1 | 12 | 0.0 | 0 | 0.0 | 2 | 0.0 |
| **15804** | T | C | 11603 | 0.0 | 17539 | 0.0 | 177 | 0.0 | 26 | 0.0 | 4053 | 0.0 | 11 | 54.6 | 0 | 0.0 | 5 | 0.0 |
| **15851** | A | C | 1267 | 1.7 | 7582 | 0.8 | 47 | 14.9 | 7 | 0.0 | 1076 | 0.0 | 3 | 0.0 | 1 | 0.0 | 1 | 0.0 |
| **15854** | T | A | 1080 | 7.0 | 7369 | 4.2 | 48 | 14.6 | 6 | 0.0 | 1001 | 0.0 | 3 | 0.0 | 1 | 0.0 | 0 | 0.0 |
| **15856** | A | G | 1300 | 5.5 | 7489 | 3.5 | 46 | 15.2 | 8 | 37.5 | 1074 | 0.0 | 4 | 0.0 | 1 | 0.0 | 1 | 0.0 |

| POS | REF | ALT | Relative of P1 | | P1.1 | | P1.2 | | P1.3 | | P1.4 | | P1.5 | | P1.6 | | P1.7 | |
|---|---|---|---|---|---|---|---|---|---|---|---|---|---|---|---|---|---|---|
| | | | N reads | Avg het | N reads | Avg het | N reads | Avg het | N reads | Avg het | N reads | Avg het | N reads | Avg het | N reads | Avg het | N reads | Avg het |
| **15982** | G | A | 14104 | 1.2 | 20818 | 1.4 | 67 | 1.5 | 372 | 0.0 | 12298 | 0.0 | 9 | 0.0 | 4 | 100.0 | 14 | 0.0 |
| **15988** | A | G | 5195 | 0.0 | 8171 | 0.0 | 16 | 0.0 | 180 | 0.0 | 5409 | 6.2 | 3 | 0.0 | 2 | 0.0 | 3 | 0.0 |
| **16006** | A | G | 15372 | 0.0 | 14953 | 0.3 | 29 | 13.8 | 484 | 0.0 | 13590 | 5.6 | 3 | 0.0 | 3 | 0.0 | 12 | 0.0 |
| **16385** | G | A | 18490 | 0.0 | 49697 | 0.0 | 444 | 0.0 | 839 | 0.0 | 5551 | 5.3 | 15 | 0.0 | 4796 | 24.9 | 24 | 0.0 |
| **16424** | T | C | 15054 | 0.6 | 46381 | 0.4 | 432 | 0.0 | 757 | 0.0 | 4983 | 0.0 | 14 | 0.0 | 4425 | 21.0 | 23 | 0.0 |
| **16548** | T | C | 10605 | 0.0 | 30434 | 0.0 | 238 | 0.0 | 49 | 0.0 | 5393 | 0.0 | 8 | 0.0 | 305 | 70.2 | 6 | 0.0 |
| **16801** | A | T | 6649 | 0.0 | 20543 | 0.0 | 70 | 0.0 | 21 | 0.0 | 7907 | 13.6 | 6 | 0.0 | 255 | 0.0 | 4 | 0.0 |
| **17121** | A | G | 1798 | 0.0 | 14184 | 0.0 | 9 | 0.0 | 34 | 0.0 | 1384 | 0.0 | 6 | 0.0 | 320 | 100.0 | 64 | 0.0 |
| **17164** | T | C | 24748 | 0.0 | 47086 | 0.2 | 681 | 0.0 | 967 | 0.0 | 35927 | 0.0 | 9 | 0.0 | 357 | 0.0 | 63 | 52.4 |
| **17335** | A | G | 4089 | 0.0 | 19583 | 0.0 | 12 | 0.0 | 112 | 0.0 | 2428 | 0.0 | 34 | 11.8 | 1 | 0.0 | 6 | 0.0 |
| **17483** | C | T | 5757 | 0.0 | 10435 | 0.0 | 277 | 0.0 | 51 | 0.0 | 2759 | 6.3 | 5 | 0.0 | 144 | 0.0 | 5 | 0.0 |
| **17485** | C | T | 5685 | 0.0 | 10459 | 0.0 | 269 | 0.0 | 53 | 0.0 | 2731 | 6.1 | 5 | 0.0 | 143 | 0.0 | 4 | 0.0 |
| **17927** | C | T | 7383 | 0.0 | 23691 | 0.0 | 115 | 0.0 | 23 | 0.0 | 5157 | 6.1 | 7 | 0.0 | 0 | 0.0 | 6 | 0.0 |
| **18342** | T | C | 8198 | 0.0 | 28712 | 0.0 | 373 | 0.0 | 31 | 71.0 | 5169 | 0.0 | 6 | 0.0 | 214 | 0.0 | 2 | 0.0 |
| **18688** | T | C | 523 | 0.0 | 7635 | 0.7 | 210 | 6.7 | 5 | 0.0 | 3 | 0.0 | 1 | 0.0 | 1 | 0.0 | 1 | 0.0 |
| **18747** | C | T | 3583 | 0.0 | 16366 | 0.0 | 383 | 0.0 | 464 | 0.0 | 4919 | 26.2 | 3 | 0.0 | 66 | 0.0 | 68 | 0.0 |
| **19087** | T | C | 6952 | 0.0 | 23103 | 0.0 | 24 | 0.0 | 21 | 0.0 | 4127 | 0.0 | 4 | 0.0 | 1257 | 0.0 | 41 | 34.2 |
| **19287** | T | C | 8400 | 0.0 | 13344 | 0.0 | 170 | 0.0 | 13 | 0.0 | 2963 | 7.3 | 2 | 0.0 | 131 | 0.0 | 6 | 0.0 |
| **19521** | T | C | 7676 | 0.0 | 20076 | 0.0 | 130 | 0.0 | 37 | 0.0 | 1737 | 42.0 | 6 | 0.0 | 1 | 0.0 | 9 | 0.0 |
| **19819** | C | T | 4254 | 0.0 | 13493 | 0.0 | 209 | 6.2 | 21 | 0.0 | 2727 | 0.0 | 6 | 0.0 | 199 | 0.0 | 3 | 0.0 |
| **20262** | A | G | 334 | 6.6 | 8613 | 0.0 | 52 | 0.0 | 3 | 0.0 | 3 | 0.0 | 8 | 0.0 | 0 | 0.0 | 3 | 0.0 |
| **20988** | T | C | 3484 | 0.0 | 17926 | 0.4 | 539 | 5.0 | 78 | 0.0 | 1878 | 0.0 | 69 | 0.0 | 0 | 0.0 | 23 | 0.0 |
| **21171** | A | G | 7670 | 0.0 | 21630 | 0.0 | 250 | 9.2 | 145 | 0.0 | 11495 | 0.0 | 3 | 0.0 | 461 | 0.0 | 4 | 0.0 |
| **21232** | G | A | 4397 | 0.0 | 22092 | 0.0 | 13 | 0.0 | 497 | 0.0 | 1860 | 21.6 | 277 | 0.0 | 696 | 0.0 | 562 | 0.0 |
| **21429** | T | A | 1078 | 0.0 | 13512 | 0.0 | 6 | 0.0 | 9 | 0.0 | 6 | 66.7 | 2 | 0.0 | 0 | 0.0 | 3 | 0.0 |
| **21765** | T | C | 5410 | 0.0 | 24104 | 0.0 | 23 | 0.0 | 24 | 0.0 | 13373 | 0.0 | 7 | 0.0 | 39 | 10.3 | 15 | 0.0 |
| **21788** | A | G | 5632 | 0.0 | 24110 | 0.0 | 23 | 26.1 | 26 | 0.0 | 13483 | 23.0 | 7 | 0.0 | 39 | 0.0 | 14 | 0.0 |
| **21908** | T | C | 6908 | 0.0 | 29762 | 0.0 | 24 | 0.0 | 62 | 0.0 | 2568 | 5.8 | 10 | 0.0 | 604 | 0.0 | 18 | 0.0 |
| **22025** | A | G | 1533 | 0.0 | 16354 | 0.2 | 5 | 0.0 | 30 | 0.0 | 198 | 0.0 | 5 | 0.0 | 539 | 43.6 | 7 | 0.0 |
| **22396** | A | G | 4378 | 0.0 | 13868 | 0.0 | 195 | 0.0 | 34 | 0.0 | 1502 | 15.5 | 1 | 0.0 | 180 | 0.0 | 24 | 0.0 |
| **23086** | C | T | 5537 | 5.1 | 11337 | 0.0 | 9 | 0.0 | 31 | 0.0 | 3556 | 0.0 | 4 | 0.0 | 146 | 0.0 | 8 | 0.0 |
| **23163** | T | C | 5578 | 0.0 | 19864 | 0.2 | 191 | 0.0 | 26 | 0.0 | 2817 | 0.0 | 8 | 0.0 | 451 | 0.0 | 15 | 46.7 |
| **23403** | A | G | 6427 | 100.0 | 36321 | 100.0 | 26 | 100.0 | 359 | 100.0 | 12792 | 100.0 | 6 | 100.0 | 475 | 100.0 | 17 | 100.0 |
| **23693** | T | C | 7840 | 0.0 | 38343 | 0.2 | 27 | 0.0 | 419 | 0.0 | 6470 | 0.4 | 109 | 0.0 | 5122 | 0.0 | 27 | 14.8 |
| **23698** | T | C | 6269 | 0.0 | 36625 | 0.0 | 24 | 0.0 | 316 | 0.0 | 6112 | 0.0 | 84 | 0.0 | 4220 | 22.9 | 22 | 0.0 |
| **23704** | A | G | 7788 | 0.0 | 37152 | 0.0 | 28 | 0.0 | 390 | 0.0 | 6443 | 0.0 | 97 | 0.0 | 4742 | 17.0 | 27 | 0.0 |
| **23884** | A | G | 2928 | 0.0 | 18161 | 0.0 | 6 | 0.0 | 18 | 0.0 | 1071 | 7.8 | 5 | 0.0 | 0 | 0.0 | 6 | 0.0 |
| **24077** | G | T | 5387 | 100.0 | 25846 | 100.0 | 36 | 16.7 | 159 | 0.0 | 15747 | 7.3 | 7 | 0.0 | 694 | 0.0 | 17 | 0.0 |
| **24104** | G | A | 5552 | 0.0 | 26157 | 0.0 | 34 | 0.0 | 162 | 0.0 | 15723 | 0.0 | 7 | 0.0 | 698 | 0.0 | 18 | 22.2 |
| **24161** | G | A | 6292 | 0.0 | 26151 | 0.0 | 25 | 0.0 | 75 | 0.0 | 7779 | 0.4 | 3 | 0.0 | 392 | 37.2 | 19 | 0.0 |
| **24447** | T | C | 7455 | 0.0 | 22767 | 0.0 | 30 | 0.0 | 620 | 0.0 | 14751 | 0.0 | 9 | 0.0 | 933 | 54.5 | 10 | 0.0 |

| POS | REF | ALT | Relative of P1 | | P1.1 | | P1.2 | | P1.3 | | P1.4 | | P1.5 | | P1.6 | | P1.7 | |
|---|---|---|---|---|---|---|---|---|---|---|---|---|---|---|---|---|---|---|
| | | | **N reads** | **Avg het** | **N reads** | **Avg het** | **N reads** | **Avg het** | **N reads** | **Avg het** | **N reads** | **Avg het** | **N reads** | **Avg het** | **N reads** | **Avg het** | **N reads** | **Avg het** |
| **24858** | G | A | 4626 | 0.0 | 25986 | 0.0 | 320 | 0.0 | 35 | 0.0 | 3529 | 6.9 | 15 | 0.0 | 428 | 0.0 | 1 | 0.0 |
| **24873** | T | C | 5507 | 0.0 | 25568 | 0.0 | 352 | 0.0 | 42 | 0.0 | 3816 | 6.7 | 15 | 0.0 | 474 | 0.0 | 1 | 0.0 |
| **24993** | A | G | 5618 | 0.0 | 27328 | 0.0 | 340 | 0.0 | 44 | 0.0 | 4369 | 0.0 | 12 | 0.0 | 466 | 33.7 | 2 | 0.0 |
| **25046** | C | T | 6197 | 0.0 | 28442 | 0.0 | 341 | 0.0 | 46 | 0.0 | 4588 | 18.2 | 11 | 0.0 | 477 | 0.0 | 4 | 0.0 |
| **25105** | A | G | 1001 | 0.0 | 6695 | 0.0 | 2 | 0.0 | 9 | 0.0 | 575 | 7.1 | 0 | 0.0 | 0 | 0.0 | 0 | 0.0 |
| **25440** | G | A | 10726 | 0.0 | 36120 | 0.0 | 17 | 0.0 | 260 | 0.0 | 7624 | 2.7 | 12 | 0.0 | 355 | 80.9 | 16 | 0.0 |
| **25556** | T | C | 10491 | 0.0 | 35154 | 0.3 | 447 | 0.0 | 1068 | 0.0 | 6698 | 7.7 | 25 | 0.0 | 3137 | 0.0 | 1087 | 12.5 |
| **25608** | A | G | 12114 | 0.0 | 37929 | 0.2 | 894 | 49.2 | 2183 | 0.0 | 3470 | 0.0 | 35 | 0.0 | 5303 | 0.0 | 1878 | 0.0 |
| **25657** | A | G | 11923 | 0.0 | 23571 | 0.0 | 52 | 0.0 | 77 | 0.0 | 13607 | 8.5 | 4 | 0.0 | 3 | 0.0 | 7 | 0.0 |
| **25677** | G | A | 14040 | 0.0 | 31970 | 0.0 | 56 | 0.0 | 81 | 0.0 | 12091 | 13.8 | 13 | 0.0 | 3 | 0.0 | 9 | 0.0 |
| **25785** | G | A | 9363 | 0.0 | 31450 | 0.0 | 27 | 0.0 | 53 | 0.0 | 9595 | 14.9 | 6 | 0.0 | 1 | 0.0 | 6 | 0.0 |
| **25789** | T | C | 11930 | 0.0 | 32893 | 0.0 | 37 | 0.0 | 71 | 0.0 | 10905 | 6.3 | 12 | 0.0 | 1 | 0.0 | 8 | 0.0 |
| **25892** | T | C | 5982 | 0.0 | 14331 | 0.0 | 15 | 0.0 | 99 | 0.0 | 4814 | 5.7 | 3 | 0.0 | 424 | 0.0 | 5 | 0.0 |
| **25934** | A | G | 12216 | 0.0 | 39205 | 0.0 | 997 | 0.0 | 284 | 0.0 | 4682 | 5.8 | 6 | 0.0 | 1559 | 0.0 | 102 | 0.0 |
| **26019** | A | G | 14574 | 0.0 | 40144 | 0.0 | 1174 | 0.0 | 336 | 0.0 | 4907 | 5.5 | 6 | 0.0 | 1848 | 0.0 | 145 | 0.0 |
| **26788** | G | A | 2984 | 0.0 | 17121 | 0.0 | 4 | 0.0 | 9 | 0.0 | 999 | 10.7 | 8 | 0.0 | 0 | 0.0 | 6 | 0.0 |
| **26895** | C | T | 10907 | 0.0 | 27539 | 0.0 | 29 | 24.1 | 277 | 0.0 | 8913 | 32.6 | 9 | 0.0 | 0 | 0.0 | 20 | 0.0 |
| **27014** | G | A | 10617 | 0.0 | 27763 | 0.0 | 28 | 25.0 | 262 | 0.0 | 8755 | 0.0 | 9 | 0.0 | 0 | 0.0 | 18 | 0.0 |
| **27085** | C | T | 9397 | 0.0 | 23364 | 0.0 | 65 | 0.0 | 2096 | 0.0 | 26165 | 0.0 | 18 | 0.0 | 2 | 100.0 | 20 | 0.0 |
| **27494** | C | T | 1839 | 0.0 | 14224 | 6.7 | 3 | 0.0 | 43 | 0.0 | 1253 | 0.0 | 4 | 0.0 | 6 | 0.0 | 5 | 0.0 |
| **27614** | T | C | 13140 | 0.0 | 20306 | 0.0 | 13 | 0.0 | 29 | 0.0 | 4163 | 9.6 | 7 | 0.0 | 413 | 0.0 | 6 | 0.0 |
| **27652** | T | C | 13381 | 0.0 | 25860 | 0.0 | 47 | 0.0 | 29 | 0.0 | 9013 | 0.0 | 6 | 0.0 | 1068 | 45.6 | 7 | 0.0 |
| **28494** | T | C | 1854 | 0.0 | 10932 | 0.0 | 145 | 11.0 | 24 | 0.0 | 1 | 0.0 | 3 | 0.0 | 154 | 0.0 | 2 | 0.0 |
| **28544** | A | G | 21304 | 0.0 | 35266 | 0.0 | 170 | 0.0 | 67 | 0.0 | 2874 | 6.9 | 9 | 0.0 | 170 | 0.0 | 15 | 0.0 |
| **28775** | C | T | 7224 | 0.0 | 31149 | 0.0 | 4 | 0.0 | 144 | 0.0 | 95 | 0.0 | 8 | 0.0 | 807 | 45.0 | 7 | 0.0 |
| **28881** | G | A | 8982 | 0.0 | 15939 | 0.0 | 150 | 3.3 | 35 | 100.0 | 2569 | 100.0 | 17 | 100.0 | 1 | 100.0 | 6 | 83.3 |
| **28882** | G | A | 15071 | 0.0 | 22435 | 0.0 | 209 | 2.9 | 24 | 100.0 | 2110 | 100.0 | 10 | 100.0 | 0 | 0.0 | 4 | 75.0 |
| **28883** | G | C | 15084 | 0.0 | 22428 | 0.0 | 208 | 2.4 | 34 | 100.0 | 2533 | 100.0 | 15 | 100.0 | 1 | 100.0 | 6 | 83.3 |
| **28916** | G | A | 13707 | 0.0 | 27841 | 0.0 | 201 | 0.0 | 175 | 7.4 | 3988 | 0.0 | 34 | 0.0 | 105 | 0.0 | 88 | 0.0 |
| **28933** | T | C | 11432 | 0.8 | 27048 | 0.4 | 175 | 0.0 | 140 | 0.0 | 3486 | 0.0 | 27 | 0.0 | 99 | 56.6 | 80 | 0.0 |
| **29061** | C | T | 8528 | 0.0 | 21694 | 0.0 | 8 | 0.0 | 5 | 0.0 | 266 | 10.5 | 95 | 0.0 | 12 | 0.0 | 7 | 0.0 |
| **29314** | A | G | 5754 | 0.0 | 25412 | 0.2 | 117 | 0.0 | 23 | 0.0 | 924 | 0.0 | 8 | 0.0 | 107 | 7.5 | 9 | 0.0 |
| **29555** | C | T | 2937 | 100.0 | 17539 | 100.0 | 6 | 100.0 | 11 | 0.0 | 0 | 0.0 | 12 | 0.0 | 0 | 0.0 | 7 | 0.0 |
| **29609** | T | C | 6513 | 0.0 | 32394 | 0.0 | 213 | 0.0 | 141 | 0.0 | 7676 | 5.0 | 14 | 0.0 | 203 | 0.0 | 9 | 0.0 |
| **29775** | T | C | 6349 | 0.0 | 25897 | 0.0 | 242 | 0.0 | 394 | 0.0 | 10204 | 2.5 | 133 | 0.0 | 703 | 51.2 | 298 | 0.0 |

**Supplementary Table S5.** List of the significantly associated variants used for PRS calculation based on the A2_ALL dataset of "very severe respiratory confirmed covid" (n=2,972) vs. population (n=284,472).

| CHR | POS | REF | ALT | rsid | Meta Effect Size | Meta P-value | Meta AF | Portuguese population Cohort AF |
|---|---|---|---|---|---|---|---|---|
| **1** | 26504654 | G | A | rs185041400 | 4.8613 | 3.55E-06 | 0.001845 | 0.00196 |
| **1** | 44377503 | G | A | rs4314918 | -0.19251 | 1.64E-06 | 0.7941 | 0.76932 |
| **1** | 112764378 | C | T | rs2919285 | -0.19459 | 2.15E-07 | 0.8032 | 0.78099 |
| **1** | 112785942 | C | G | rs7538140 | 0.16488 | 9.57E-06 | 0.2219 | 0.21499 |
| **1** | 203954605 | T | C | rs188564545 | 2.4201 | 4.13E-06 | 0.00984 | 0.00158 |
| **1** | 231996139 | A | C | rs75711735 | 0.58372 | 5.52E-06 | 0.0374 | 0.02181 |
| **2** | 43251821 | G | A | rs56186825 | 1.6652 | 3.27E-06 | 0.008928 | 0.00517 |
| **2** | 130431716 | C | T | rs10210034 | -0.14541 | 5.68E-06 | 0.5131 | 0.48767 |
| **2** | 137341234 | A | T | rs531390248 | 3.36 | 7.77E-06 | 0.002369 | 0.00016 |
| **2** | 217725954 | C | T | rs6722107 | 0.90977 | 7.93E-06 | 0.008955 | 0.01814 |
| **3** | 38579153 | A | G | rs112661205 | 2.2515 | 8.46E-06 | 0.003901 | 0.00245 |
| **3** | 39204989 | C | T | rs148889878 | 1.3637 | 1.70E-06 | 0.00486 | 0.00941 |
| **3** | 45889921 | A | T | rs35081325 | 0.70561 | 3.82E-39 | 0.09694 | 0.05894 |
| **3** | 45908859 | G | A | rs75826707 | 0.71416 | 4.54E-13 | 0.04609 | 0.0089 |
| **3** | 46032388 | G | C | NA | -0.18152 | 9.03E-08 | 0.6364 | 0.68145 |
| **3** | 46049765 | T | C | rs13433997 | 0.42141 | 3.66E-20 | 0.1564 | 0.10061 |
| **3** | 46119791 | T | C | rs13434336 | 0.20352 | 1.81E-09 | 0.3334 | 0.39426 |
| **3** | 46222037 | A | G | rs115102354 | 0.50009 | 3.25E-13 | 0.07706 | 0.02963 |
| **3** | 46227171 | T | G | rs13062450 | 0.28632 | 6.78E-08 | 0.1518 | 0.08022 |
| **3** | 46306474 | T | C | rs7631853 | 0.34086 | 2.20E-09 | 0.1289 | 0.06356 |
| **3** | 69056121 | G | A | rs76821671 | 0.24803 | 5.41E-06 | 0.08798 | 0.07825 |
| **3** | 112914296 | A | G | rs182721950 | 1.8166 | 6.32E-06 | 0.001878 | 0.01382 |
| **3** | 143251664 | A | G | rs62269771 | 0.33933 | 7.46E-06 | 0.08874 | 0.1556 |
| **3** | 145864511 | G | A | rs965032 | -0.16202 | 2.04E-06 | 0.6111 | 0.65497 |
| **3** | 159270026 | G | C | rs113427422 | 0.48165 | 2.85E-06 | 0.0559 | 0.0693 |
| **3** | 165154092 | C | A | NA | -0.14427 | 7.20E-06 | 0.4587 | 0.40861 |
| **4** | 25475602 | A | G | rs4697099 | -0.23761 | 8.97E-06 | 0.1697 | 0.12552 |
| **4** | 36530269 | C | G | rs61796478 | 1.5601 | 2.62E-06 | 0.008746 | 0.01632 |
| **4** | 122910410 | G | A | rs7686809 | -0.50162 | 1.74E-07 | 0.9453 | 0.88729 |
| **5** | 7505930 | C | T | rs192229354 | 3.5975 | 4.20E-06 | 0.002109 | 0.00484 |
| **5** | 17130850 | T | C | rs790210 | 0.54784 | 5.30E-06 | 0.03515 | 5.00E-05 |
| **5** | 17142133 | G | T | rs645922 | 0.58518 | 4.78E-06 | 0.03495 | 0.02149 |
| **5** | 24843084 | A | G | rs141962254 | 2.9487 | 6.74E-06 | 0.001148 | 0.0015 |
| **5** | 58786782 | G | T | rs146410305 | 0.67245 | 2.24E-06 | 0.02408 | 0.0062 |
| **5** | 79070021 | C | T | rs114128029 | 1.1891 | 1.60E-06 | 0.01538 | 0.00427 |
| **5** | 131740656 | A | C | rs13168774 | -0.19091 | 6.61E-06 | 0.8634 | 0.83698 |
| **5** | 154604690 | C | G | NA | 0.2305 | 8.22E-06 | 0.1513 | 0.07799 |
| **5** | 164850943 | G | A | rs6880269 | -0.2311 | 4.99E-06 | 0.5075 | 0.50703 |
| **6** | 7549628 | C | T | rs2299036 | 0.29664 | 9.19E-07 | 0.1795 | 0.17953 |
| **6** | 31121426 | G | A | rs143334143 | 0.45345 | 5.60E-17 | 0.1437 | 0.12398 |
| **6** | 32667171 | A | T | rs1794280 | -0.30549 | 1.71E-08 | 0.1002 | 0.09167 |
| **6** | 33055355 | A | G | NA | 0.27651 | 2.76E-07 | 0.07587 | 0.09132 |
| **6** | 41719110 | T | C | NA | -0.56103 | 7.10E-06 | 0.976 | 0.96753 |
| **6** | 88576981 | T | C | rs6935448 | -0.22002 | 5.42E-06 | 0.8219 | 0.83752 |
| **6** | 98715160 | G | A | rs117937941 | 0.47232 | 2.25E-06 | 0.005898 | 0.04364 |
| **7** | 37831003 | A | G | rs183729083 | 1.8366 | 9.03E-06 | 0.005809 | 0.00277 |
| **7** | 54647894 | A | C | rs622568 | 0.27338 | 2.83E-10 | 0.1608 | 0.07366 |
| **7** | 88639135 | T | C | rs78211246 | -0.19717 | 6.19E-06 | 0.1759 | 0.17091 |
| **7** | 107607902 | C | T | rs2237698 | 0.22516 | 6.99E-06 | 0.09765 | 0.1192 |
| **7** | 113317708 | T | C | rs12705891 | 0.16763 | 6.31E-07 | 0.3918 | 0.41102 |
| **8** | 1779298 | C | T | rs180717749 | 0.69669 | 4.98E-06 | 0.002375 | 0.00724 |
| **8** | 71700365 | C | T | rs147667474 | 2.0373 | 2.89E-06 | 0.01164 | 0.00431 |
| **8** | 141857912 | A | G | rs149938155 | 0.43195 | 5.95E-06 | 0.01057 | 0.0238 |
| **9** | 123550027 | G | A | NA | 0.28206 | 9.25E-06 | 0.7073 | 0.7653 |
| **10** | 33178246 | A | C | NA | -0.21564 | 6.72E-06 | 0.8609 | 0.86629 |
| **10** | 44340072 | T | A | rs118052809 | 0.55026 | 2.60E-06 | 0.01048 | 0.01291 |
| **10** | 97252761 | A | T | rs117098321 | 1.0297 | 3.69E-07 | 0.008664 | 0.01872 |
| **11** | 22828273 | C | T | rs78594643 | -0.54047 | 9.33E-06 | 0.01995 | 0.02303 |
| **11** | 35402078 | T | C | rs1923302 | -0.17211 | 9.34E-06 | 0.2239 | 0.21984 |
| **11** | 44727667 | A | G | rs4450162 | 0.46666 | 3.97E-06 | 0.04025 | 0.06024 |
| **11** | 125792416 | T | C | rs662722 | -0.24615 | 5.55E-06 | 0.7416 | 0.67452 |

| CHR | POS | REF | ALT | rsid | Meta Effect Size | Meta P-value | Meta AF | Portuguese population Cohort AF |
|---|---|---|---|---|---|---|---|---|
| **12** | 29229856 | G | A | NA | -0.19661 | 2.75E-06 | 0.8177 | 0.81272 |
| **12** | 103014757 | C | A | NA | -0.37565 | 1.98E-14 | 0.8746 | 0.86556 |
| **12** | 113381956 | C | T | rs2269899 | 0.21454 | 8.55E-10 | 0.72 | 0.62316 |
| **12** | 113385375 | T | C | rs10850104 | 0.19459 | 2.36E-07 | 0.3694 | 0.19024 |
| **13** | 44342409 | A | G | rs9533610 | -0.20652 | 7.76E-06 | 0.8439 | 0.81248 |
| **13** | 44604709 | C | G | NA | 0.47437 | 2.01E-06 | 0.007279 | 0.02136 |
| **13** | 67940439 | A | T | rs9592514 | 2.5513 | 5.71E-06 | 0.00228 | 0.00378 |
| **14** | 81096699 | C | T | rs8016670 | -0.27455 | 3.96E-06 | 0.9602 | 0.91986 |
| **15** | 37079890 | A | G | rs149402468 | 1.9556 | 6.47E-06 | 0.003407 | 0.0086 |
| **15** | 79766794 | G | A | NA | -0.1936 | 1.42E-06 | 0.2287 | 0.23173 |
| **15** | 99973286 | T | G | rs74035732 | -0.35901 | 7.65E-06 | 0.04311 | 0.05008 |
| **17** | 10101496 | C | T | rs149399480 | 1.8031 | 8.46E-07 | 0.006129 | 0.00804 |
| **17** | 17462512 | C | T | rs568997551 | 2.2931 | 3.45E-06 | 0.01207 | 0.00343 |
| **17** | 33976296 | C | T | NA | -0.18834 | 7.91E-06 | 0.7955 | 0.80944 |
| **17** | 76252183 | G | A | NA | 0.36136 | 2.62E-06 | 0.1979 | 0.14406 |
| **18** | 36300243 | A | G | rs142354687 | 3.8192 | 5.03E-06 | 0.002618 | 0.00375 |
| **19** | 4723670 | C | A | NA | 0.25516 | 6.69E-13 | 0.3364 | 0.27724 |
| **19** | 10427721 | T | A | NA | 0.38141 | 1.31E-07 | 0.03602 | 0.03906 |
| **19** | 10466123 | C | T | rs11085727 | 0.18858 | 1.23E-07 | 0.267 | 0.30801 |
| **19** | 10596988 | C | A | rs45524632 | 0.51705 | 8.67E-07 | 0.01571 | 0.01407 |
| **19** | 17505834 | G | C | NA | 0.30266 | 3.32E-06 | 0.7688 | 0.82365 |
| **19** | 43266536 | C | G | rs112599803 | 0.66574 | 2.48E-06 | 0.06424 | 0.04464 |
| **19** | 43905258 | C | T | rs75028208 | 1.3477 | 2.23E-06 | 0.01459 | 0.03103 |
| **19** | 56456647 | G | A | rs141789023 | 0.56968 | 2.89E-06 | 0.1156 | 0.03703 |
| **20** | 24479907 | C | T | rs547278572 | 2.8651 | 5.12E-06 | 0.002402 | 0.00149 |
| **20** | 32781699 | G | C | rs546079703 | 2.4138 | 1.72E-07 | 0.004147 | 0.00014 |
| **20** | 37563008 | T | C | rs186431128 | 2.7074 | 5.59E-06 | 0.002141 | 0.00028 |
| **20** | 57549023 | C | T | rs150240336 | 1.4451 | 5.30E-06 | 0.0143 | 0.01093 |
| **21** | 34615210 | T | C | rs13050728 | -0.19739 | 1.84E-08 | 0.6114 | 0.69188 |
| **22** | 41252291 | T | C | rs192261735 | 0.63286 | 3.12E-06 | 0.006277 | 0.0072 |

**Supplementary Table S6.** List of the significantly associated variants used for PRS calculation based on the B2_ALL dataset of "hospitalised covid" (n=6,492) vs. population (n=1,012,809).

| CHR | POS | REF | ALT | rsid | Meta Effect Size | Meta P-value | Meta AF | Portuguese population Cohort AF |
|---|---|---|---|---|---|---|---|---|
| **1** | 53771860 | G | A | rs115038483 | -0.42964 | 6.43E-06 | 0.01405 | 0.01063 |
| **1** | 65449821 | G | A | rs4454580 | 0.18513 | 2.22E-06 | 0.1157 | 0.13099 |
| **1** | 91208514 | A | C | rs2166172 | 0.10713 | 8.63E-06 | 0.4079 | 0.35832 |
| **1** | 237277098 | A | C | rs9287218 | -0.24463 | 2.82E-06 | 0.05047 | 0.04674 |
| **2** | 162936216 | C | T | rs117888248 | 0.6907 | 2.37E-06 | 0.01038 | 0.02342 |
| **2** | 182809457 | C | G | rs74799459 | 0.30726 | 9.18E-06 | 0.03178 | 0.04498 |
| **2** | 195035942 | A | G | rs62186769 | 0.61202 | 8.87E-06 | 0.008733 | 0.00035 |
| **3** | 7794348 | C | T | NA | -0.14468 | 9.44E-06 | 0.2636 | 0.24427 |
| **3** | 27526516 | C | T | rs6771541 | 0.14359 | 1.17E-06 | 0.3799 | 0.44167 |
| **3** | 45553090 | G | A | rs79939301 | 0.25641 | 5.57E-07 | 0.08346 | 0.02906 |
| **3** | 45798226 | C | T | rs17213127 | 0.38764 | 7.66E-08 | 0.04395 | 0.0246 |
| **3** | 45818880 | G | C | NA | 0.21697 | 3.02E-06 | 0.1237 | 0.11258 |
| **3** | 45822010 | T | C | rs73062378 | 0.18408 | 8.04E-06 | 0.2093 | 0.15026 |
| **3** | 45889921 | A | T | rs35081325 | 0.59959 | 9.52E-50 | 0.08054 | 0.05894 |
| **3** | 45908859 | G | A | rs75826707 | 0.64829 | 3.59E-16 | 0.03148 | 0.0089 |
| **3** | 45910870 | G | A | rs2191031 | 0.21 | 2.31E-09 | 0.2034 | 0.18051 |
| **3** | 46042413 | A | T | NA | -0.15672 | 7.31E-11 | 0.5993 | 0.66127 |
| **3** | 46047767 | G | C | rs4234452 | -0.10771 | 7.36E-06 | 0.4014 | 0.38653 |
| **3** | 46049765 | T | C | rs13433997 | 0.37339 | 1.66E-29 | 0.1322 | 0.10061 |
| **3** | 46093858 | G | A | rs13098271 | 0.15548 | 2.01E-10 | 0.3283 | 0.37582 |
| **3** | 46194589 | C | A | rs71327036 | 0.28439 | 8.85E-14 | 0.1052 | 0.06948 |
| **3** | 46222037 | A | G | rs115102354 | 0.45004 | 1.97E-18 | 0.06527 | 0.02963 |
| **3** | 46301423 | T | A | rs11919884 | 0.27677 | 5.95E-12 | 0.09721 | 0.07677 |
| **3** | 46464017 | A | C | rs34671664 | 0.19128 | 3.12E-07 | 0.1241 | 0.06349 |
| **4** | 36530269 | C | G | rs61796478 | 1.0631 | 1.66E-06 | 0.005496 | 0.01632 |

| CHR | POS | REF | ALT | rsid | Meta Effect Size | Meta P-value | Meta AF | Portuguese population Cohort AF |
|---|---|---|---|---|---|---|---|---|
| **4** | 142430667 | A | T | NA | 0.4692 | 1.54E-06 | 0.02485 | 0.02312 |
| **5** | 56485892 | G | A | rs55737726 | 0.59065 | 3.43E-06 | 0.01254 | 0.00438 |
| **5** | 58786782 | G | T | rs146410305 | 0.54762 | 1.08E-06 | 0.01726 | 0.0062 |
| **5** | 65066483 | C | T | rs114969787 | 0.26622 | 6.51E-06 | 0.03846 | 0.06299 |
| **5** | 71698763 | T | C | rs187920931 | 1.857 | 1.43E-06 | 0.004279 | 0.00035 |
| **5** | 71763420 | A | G | rs139478596 | 2.0038 | 5.83E-07 | 0.004027 | 5.10E-04 |
| **5** | 71893271 | G | A | rs191971874 | 2.0461 | 7.90E-07 | 0.003874 | 0.00239 |
| **5** | 131740656 | A | C | rs13168774 | -0.14334 | 2.62E-06 | 0.8344 | 0.83698 |
| **5** | 162727453 | C | T | rs79833209 | 0.37801 | 2.64E-06 | 0.02819 | 0.01345 |
| **6** | 31121426 | G | A | rs143334143 | 0.2646 | 3.45E-11 | 0.08689 | 0.12398 |
| **6** | 33055355 | A | G | NA | 0.18117 | 3.58E-06 | 0.08416 | 0.09132 |
| **6** | 41497035 | C | A | NA | 0.26593 | 8.86E-10 | 0.1251 | 0.09466 |
| **6** | 41501834 | G | A | rs12175265 | 0.43782 | 4.55E-09 | 0.03502 | 0.01744 |
| **6** | 108279068 | G | A | rs200955319 | 1.0422 | 6.61E-07 | 0.006912 | 0.00534 |
| **6** | 157867030 | C | T | rs9364501 | -0.12713 | 6.96E-06 | 0.4899 | 0.45094 |
| **7** | 53493705 | G | A | rs12718791 | -0.17468 | 3.32E-06 | 0.7783 | 0.84959 |
| **7** | 54647894 | A | C | rs622568 | 0.16634 | 2.77E-07 | 0.1477 | 0.07366 |
| **7** | 107607902 | C | T | rs2237698 | 0.16851 | 2.69E-06 | 0.09467 | 0.1192 |
| **8** | 9875204 | T | C | rs1396186 | 0.16268 | 2.64E-06 | 0.2264 | 0.20498 |
| **8** | 10010765 | G | C | NA | 0.15341 | 7.41E-06 | 0.1962 | 0.18969 |
| **8** | 15343283 | T | C | rs114717629 | 1.1699 | 9.23E-06 | 0.00655 | 0.00713 |
| **8** | 59903271 | T | C | rs7823862 | -0.20454 | 1.65E-06 | 0.06634 | 0.08014 |
| **8** | 110059637 | A | C | rs189378134 | 1.0541 | 7.28E-07 | 0.007647 | 0.0043 |
| **8** | 110339840 | T | A | rs144837205 | 0.96534 | 3.58E-06 | 0.007712 | 0.00424 |
| **9** | 15397969 | A | C | rs2798716 | -0.18197 | 6.51E-06 | 0.9302 | 0.87977 |
| **9** | 26974745 | C | T | rs150788916 | 1.4433 | 3.49E-06 | 0.004252 | 0.00346 |
| **10** | 12667474 | C | G | rs4310517 | -0.1396 | 6.65E-06 | 0.7521 | 0.6526 |
| **10** | 20642449 | T | C | NA | 1.5167 | 6.14E-06 | 0.004527 | 0.00035 |
| **10** | 44340072 | T | A | rs118052809 | 0.45242 | 1.31E-06 | 0.02222 | 0.01291 |
| **10** | 70172332 | G | A | NA | 0.35143 | 5.77E-06 | 0.02673 | 0.02881 |
| **10** | 121010105 | A | C | rs112969140 | 0.20563 | 8.00E-06 | 0.08061 | 0.05773 |
| **11** | 1005506 | A | G | rs147685259 | 1.2734 | 4.82E-06 | 0.006075 | 0.00526 |
| **11** | 113106528 | G | T | NA | -0.27335 | 2.90E-06 | 0.05598 | 0.04889 |
| **12** | 31392117 | C | T | rs75594480 | 0.90255 | 4.16E-06 | 0.01692 | 0.01057 |
| **12** | 103014757 | C | A | NA | -0.18231 | 5.17E-07 | 0.886 | 0.86556 |
| **12** | 113362997 | T | G | NA | 0.14763 | 5.74E-09 | 0.6768 | 0.61171 |
| **12** | 129582079 | T | C | rs116993182 | 0.33965 | 1.80E-06 | 0.01796 | 0.02046 |
| **13** | 73889735 | T | C | rs2325521 | 0.28856 | 9.99E-06 | 0.03812 | 0.01865 |
| **14** | 38848003 | A | G | rs1754680 | -0.11565 | 1.90E-06 | 0.3974 | 0.39369 |
| **15** | 26780764 | T | G | NA | 0.49562 | 5.17E-06 | 0.04097 | 0.01152 |
| **15** | 55197554 | G | T | NA | 1.1663 | 9.95E-06 | 0.009457 | 0.00387 |
| **15** | 95579943 | A | G | rs145572293 | 0.61208 | 2.99E-06 | 0.01468 | 0.01191 |
| **17** | 2846004 | G | A | rs55818593 | 0.18055 | 1.50E-06 | 0.3504 | 0.33101 |
| **17** | 61954669 | G | A | rs145032579 | 0.65208 | 6.12E-06 | 0.009317 | 0.00884 |
| **18** | 25179662 | A | G | rs146116110 | 0.69167 | 3.65E-06 | 0.01124 | 0.00729 |
| **19** | 4723670 | C | A | NA | 0.18122 | 2.31E-12 | 0.315 | 0.27724 |
| **19** | 10423815 | G | A | rs8101195 | -0.13932 | 7.99E-06 | 0.8307 | 0.80739 |
| **19** | 10427721 | T | A | NA | 0.32939 | 4.50E-09 | 0.0506 | 0.03906 |
| **19** | 10596988 | C | A | rs45524632 | 0.47598 | 4.62E-09 | 0.02362 | 0.01407 |
| **21** | 25507072 | G | C | rs190545393 | 1.4221 | 4.45E-06 | 0.004305 | 0.00712 |
| **21** | 34610487 | T | C | rs1131964 | 0.13008 | 5.50E-08 | 0.5761 | 0.51179 |
| **21** | 34615210 | T | C | rs13050728 | -0.17812 | 8.83E-13 | 0.6515 | 0.69188 |
| **21** | 34620801 | A | G | rs2073362 | 0.2271 | 3.81E-08 | 0.08026 | 0.07966 |
| **22** | 19568533 | C | T | rs9604980 | 0.18951 | 2.19E-07 | 0.1053 | 0.07951 |

**Supplementary Table S7.** PRS score values for the “very severe respiratory confirmed covid” and “hospitalised covid” phenotypes in the patient and individuals from the Portuguese cohort.

| ID | “very severe respiratory confirmed covid” | “hospitalised covid” |
|---|---|---|
| **Patient** | -4.87655 | -0.03313 |
| **Control_1** | -4.90E+00 | -1.97551 |
| **Control_2** | -5.16E+00 | -1.89236 |
| **Control_3** | -7.15E+00 | -0.54649 |
| **Control_4** | -4.89E+00 | -1.71186 |
| **Control_5** | -6.21E+00 | -0.68627 |
| **Control_6** | -3.59E+00 | -1.72808 |
| **Control_7** | 2.68E-01 | 1.37396 |
| **Control_8** | -5.00E+00 | -0.4512 |
| **Control_9** | -3.28E+00 | -0.24699 |
| **Control_10** | -5.32E+00 | -1.14481 |
| **Control_11** | -4.04E+00 | -1.56374 |
| **Control_12** | -2.56E+00 | -0.64326 |
| **Control_13** | -4.67E-01 | 0.62717 |
| **Control_14** | -5.08E+00 | -1.36083 |
| **Control_15** | -4.77E+00 | -0.51723 |
| **Control_16** | -5.29E+00 | -0.59627 |
| **Control_17** | -3.30E+00 | -0.64868 |
| **Control_18** | -4.98E+00 | 0.18003 |
| **Control_19** | -5.35E+00 | -0.09047 |
| **Control_20** | -5.00E+00 | -1.34065 |
| **Control_21** | -3.38E+00 | -0.32906 |
| **Control_22** | -2.80E+00 | -1.0804 |
| **Control_23** | -4.50E+00 | -0.59643 |
| **Control_24** | -1.03E+00 | -0.39641 |
| **Control_25** | -4.99E+00 | 1.67317 |
| **Control_26** | -4.36E+00 | -0.57461 |
| **Control_27** | -4.04E+00 | 0.22938 |
| **Control_28** | -4.70E+00 | -0.18751 |
| **Control_29** | -5.64E+00 | 0.14805 |
| **Control_30** | -4.89E+00 | 0.60239 |
| **Control_31** | -4.69E+00 | -1.2565 |
| **Control_32** | -2.93E+00 | 1.90648 |
| **Control_33** | -4.09E+00 | 0.69502 |
| **Control_34** | -6.03E+00 | -2.02994 |
| **Control_35** | -9.81E-01 | -1.1441 |
| **Control_36** | -3.89E+00 | -0.97245 |
| **Control_37** | -4.96E+00 | -0.34381 |
| **Control_38** | -5.23E+00 | -0.97005 |
| **Control_39** | -5.39E+00 | 0.94992 |
| **Control_40** | -4.90E+00 | -1.05645 |
| **Control_41** | -3.18E+00 | 0.21109 |
| **Control_42** | -5.11E-01 | 0.85563 |
| **Control_43** | -4.98E+00 | -1.45702 |
| **Control_44** | -4.27E+00 | 0.3355 |
| **Control_45** | -6.39E+00 | -1.62096 |
| **Control_46** | -3.61E+00 | -1.34278 |
| **Control_47** | -7.10E+00 | -0.84061 |
| **Control_48** | -4.59E+00 | -0.6492 |
| **Control_49** | -4.75E+00 | -2.19425 |
| **Control_50** | -2.00E+00 | 0.68209 |
| **Control_51** | -2.90E+00 | 0.75568 |
| **Control_52** | -4.35E+00 | -0.85561 |
| **Control_53** | -4.19E+00 | 0.33753 |
| **Control_54** | -3.04E+00 | -0.08448 |
| **Control_55** | -5.70E+00 | 1.603 |
| **Control_56** | -6.32E+00 | -1.5768 |
| **Control_57** | -6.29E+00 | -0.7849 |
| **Control_58** | -5.36E+00 | 0.25457 |
| **Control_59** | -5.56E+00 | -1.21327 |
| **Control_60** | -3.79E+00 | -0.07656 |
| **Control_61** | -5.41E+00 | -0.10805 |
| **Control_62** | -4.54E+00 | -0.31598 |

| ID | “very severe respiratory confirmed covid” | “hospitalised covid” |
|---|---|---|
| **Control_63** | -3.18E+00 | 1.7555 |
| **Control_64** | -4.35E+00 | -0.67707 |
| **Control_65** | -3.89E+00 | 0.66665 |
| **Control_66** | -4.08E+00 | -0.62997 |
| **Control_67** | -3.23E+00 | -0.17569 |
| **Control_68** | -4.96E+00 | -0.05921 |
| **Control_69** | -4.50E+00 | 0.30167 |
| **Control_70** | -4.09E+00 | -0.61154 |
| **Control_71** | -5.43E+00 | -1.2121 |
| **Control_72** | -1.56E+00 | 3.75514 |
| **Control_73** | -4.86E+00 | -1.09983 |
| **Control_74** | -4.61E+00 | 0.20464 |
| **Control_75** | -4.76E+00 | -1.63796 |
| **Control_76** | -3.79E+00 | 0.8922 |
| **Control_77** | -2.17E+00 | 3.44269 |
| **Control_78** | -4.49E+00 | -0.05917 |
| **Control_79** | -4.28E+00 | -0.52001 |
| **Control_80** | -3.55E+00 | -1.62611 |
| **Control_81** | -4.43E+00 | -1.15316 |
| **Control_82** | -5.01E+00 | 0.56405 |
| **Control_83** | -3.23E+00 | -1.33503 |
| **Control_84** | -1.77E+00 | 2.4039 |
| **Control_85** | -5.38E+00 | -0.09866 |
| **Control_86** | -2.50E+00 | 0.87392 |
| **Control_87** | -2.11E+00 | -0.56459 |
| **Control_88** | -1.73E+00 | 0.06622 |
| **Control_89** | -2.95E+00 | 0.2116 |
| **Control_90** | -6.24E+00 | -1.60651 |
| **Control_91** | -2.30E+00 | -1.53324 |
| **Control_92** | -6.99933 | -1.61907 |
| **Control_93** | -2.45405 | 1.48303 |
| **Control_94** | -4.34987 | 1.67316 |
| **Control_95** | -4.72626 | 0.38721 |
| **Control_96** | -4.80856 | -0.0857 |
| **Control_97** | -3.04659 | 0.82665 |
| **Control_98** | -2.38563 | 0.20145 |
| **Control_99** | -2.43129 | -1.40496 |
| **Control_100** | -5.86669 | 0.14784 |
| **Control_101** | -1.35701 | 0.68653 |
| **Control_102** | -1.23382 | -1.78844 |
| **Control_103** | -4.10605 | -0.09449 |
| **Control_104** | -0.39854 | 1.65477 |
| **Control_105** | -3.63653 | 0.12011 |
| **Control_106** | -4.61694 | -0.25435 |
| **Control_107** | -4.68681 | -0.63197 |
| **Control_108** | -3.23328 | -0.08327 |
| **Control_109** | -2.97989 | 0.92944 |
| **Control_110** | -4.43776 | -0.95831 |
| **Control_111** | -2.198 | 1.2198 |
| **Control_112** | 2.42573 | 3.37488 |
| **Control_113** | -4.404 | -0.21011 |
| **Control_114** | -2.81878 | 0.12863 |
| **Control_115** | -6.21434 | 0.26661 |
| **Control_116** | -3.78205 | -1.04036 |
| **Control_117** | -4.66823 | -0.25966 |
| **Control_118** | -4.38954 | -0.68852 |
| **Control_119** | 0.30253 | 0.48275 |
| **Control_120** | -6.90608 | -1.78801 |
| **Control_121** | -4.67019 | -1.55847 |
| **Control_122** | -5.11919 | 1.00139 |
| **Control_123** | -5.31657 | -0.20587 |
| **Control_124** | -2.34435 | -0.87753 |
| **Control_125** | -3.97191 | 0.56356 |
| **Control_126** | -5.72312 | -1.35796 |
| **Control_127** | -4.12922 | -0.98348 |
| **Control_128** | -1.64077 | -0.44908 |
| **Control_129** | -5.41002 | -1.11184 |
| **Control_130** | -0.49236 | -1.32733 |
| **Control_131** | -4.5782 | -0.15519 |

| ID | “very severe respiratory confirmed covid” | “hospitalised covid” |
|---|---|---|
| **Control_132** | -5.70871 | 0.34212 |
| **Control_133** | -4.67043 | -0.68724 |
| **Control_134** | -3.25457 | -0.45825 |
| **Control_135** | -6.27244 | -2.13391 |
| **Control_136** | -3.55693 | -0.02005 |
| **Control_137** | -2.1284 | -1.45675 |
| **Control_138** | -3.72811 | -0.22451 |
| **Control_139** | -0.47755 | -0.6321 |
| **Control_140** | -4.73245 | 0.13417 |
| **Control_141** | -5.72684 | -0.78091 |
| **Control_142** | -3.63986 | -0.04767 |
| **Control_143** | -3.32291 | -1.64979 |
| **Control_144** | -3.42449 | -0.85886 |
| **Control_145** | -4.63228 | 0.18671 |
| **Control_146** | -4.94364 | -0.26755 |
| **Control_147** | -6.41413 | -1.58766 |
| **Control_148** | -3.53471 | -0.61872 |
| **Control_149** | -5.57549 | -1.11323 |
| **Control_150** | -4.74915 | -0.87604 |
| **Control_151** | -4.09005 | 0.18534 |
| **Control_152** | -5.56207 | -1.43691 |
| **Control_153** | -4.68608 | -1.04662 |
| **Control_154** | -3.91342 | 0.93384 |
| **Control_155** | -7.52479 | -2.15656 |
| **Control_156** | -4.82207 | -1.29347 |
| **Control_157** | -3.18609 | -1.0193 |
| **Control_158** | -4.86976 | -2.02661 |
| **Control_159** | -2.87761 | -0.26275 |
| **Control_160** | -4.93477 | -0.12399 |
| **Control_161** | -3.21235 | 1.42861 |
| **Control_162** | -2.77394 | 0.4151 |
| **Control_163** | -1.9014 | 0.94125 |
| **Control_164** | -1.97826 | 0.78863 |
| **Control_165** | -4.09434 | -0.14756 |
| **Control_166** | -4.74327 | -1.01587 |
| **Control_167** | -4.39397 | -0.76348 |
| **Control_168** | -4.45865 | -0.8555 |
| **Control_169** | -3.41697 | 1.12599 |
| **Control_170** | -2.99592 | 1.23925 |
| **Control_171** | -4.68866 | 0.27114 |
| **Control_172** | -5.49796 | -0.28772 |
| **Control_173** | -4.87907 | 1.22099 |
| **Control_174** | -4.09014 | -1.58057 |
| **Control_175** | -5.75143 | -0.33986 |
| **Control_176** | -3.40585 | 0.07098 |
| **Control_177** | -5.17761 | -0.7763 |
| **Control_178** | -4.0947 | -0.68351 |
| **Control_179** | -5.09591 | 0.17281 |
| **Control_180** | -4.41746 | 1.55421 |
| **Control_181** | -6.23705 | -0.33483 |
| **Control_182** | -5.67844 | -0.31867 |
| **Control_183** | -4.6586 | -0.42976 |
| **Control_184** | -5.43925 | -0.65352 |
| **Control_185** | -3.61809 | 0.00716 |
| **Control_186** | -5.14192 | 0.20938 |
| **Control_187** | -3.33742 | -0.53662 |
| **Control_188** | -4.92869 | 0.53324 |
| **Control_189** | -3.41719 | 0.165 |
| **Control_190** | -3.15799 | -0.58069 |
| **Control_191** | -4.90514 | -0.88877 |
| **Control_192** | -4.80698 | -1.24858 |
| **Control_193** | -4.67025 | 1.04502 |
| **Control_194** | -1.87099 | 0.61465 |
| **Control_195** | -4.43074 | 0.39739 |
| **Control_196** | -3.47731 | 0.58579 |
| **Control_197** | -4.36153 | 0.31097 |
| **Control_198** | -5.80403 | -0.2157 |
